# Identification of robust mortality-associated neutrophil gene programs and cytokine responses using large-scale ARDS endotracheal aspirate scRNA-seq data

**DOI:** 10.1101/2025.08.21.25333911

**Authors:** Owen Whitley, Tim Delemarre, Miguel Reyes, Kimberly Kajihara, Hannah Little-Hooy, Cynthia Chen, Marco De Simone, Min Xu, Haridha Shivram, Jason Hackney, Diana Chang, Xia Gao, Eugene Kim, Nandhini Ramamoorthi, Rafal K. Woycicki, Shoshana Zha, Charles R. Langelier, Sharookh B. Kapadia, Carrie M. Rosenberger, Huwenbo Shi

## Abstract

Myeloid dysregulation is involved in severe Acute Respiratory Distress Syndrome (ARDS), but the mechanistic understanding is still incomplete. We analyzed endotracheal aspirate (ETA) scRNA-seq data from 40 intubated patients (both COVID-19 and non-COVID-19) in the COMET cohort, to uncover cell types and gene programs associated with ARDS mortality. In cell-type-agnostic analyses, we identified neutrophils as a critical cell type prognostic for ARDS mortality. Subsequent analyses uncovered a mortality-associated neutrophil subpopulation characterized by upregulation of IFN-ɣ response and ferroptosis genes, and downregulation of TNF-ɑ response genes. These findings were validated across independent COVID-19 clinical cohorts. Analyses of *in vitro* data further suggested that gene programs active in the mortality-associated neutrophil subpopulation could be attributable to specific cytokine responses. Finally, we confirmed that this mortality-associated neutrophil subpopulation could be induced in rodents via influenza A or SARS-CoV-2 infections, providing insights for modeling this neutrophil phenotype for therapeutic development.

## Introduction

Acute Respiratory Distress Syndrome (ARDS) is a severe and critical respiratory disorder, characterized by acute lung inflammation and injury, hypoxemia, edema, and significant decline in lung function.^1–3^ The causes of ARDS are highly heterogeneous, including sepsis, bacterial pneumonia, and viral respiratory infection.^1,3–6^ With a mortality rate of approximately 40%,^1,3,5,7^ and a high financial burden on patients,^8,9^ treatment of ARDS represents an unmet medical need in the United States. Standard of care treatments for ARDS typically include prone positioning, mechanical ventilation, and extracorporeal membrane oxygenation (ECMO).^1,6,10,11^ Additional therapeutics, including the use of corticosteroids, have also been investigated for ARDS.^12–16^ However, existing therapeutics for ARDS generally have limited efficacy on reducing ARDS mortality, due to patient heterogeneity and complex ARDS disease biology.^15,17,18^ Thus, we need to develop new therapeutics that target specific ARDS pathophysiological mechanisms in well-defined patient populations.

A key biological process involved in ARDS is dysregulated immune responses.^6,16,19–24^ Recent work analyzing both COVID-19 and non-COVID-19 ARDS have implicated myeloid dysregulation as a key driver of ARDS onset and progression.^20–22,25,26^ Most of these studies focused on monocytes and macrophages. Despite the critical role of neutrophils in ARDS,^6,27–30^ the understanding of the impact of specific neutrophil subpopulations, functions, and phenotypes on patient outcome, particularly mortality, is currently limited. This is in part because assaying gene expression in neutrophils is challenging.^31–33^ However, recent technological advancements and efforts during the COVID-19 pandemic have led to an increase in neutrophil single-cell RNA-seq (scRNA-seq) studies focused on ARDS.^23,34,35^

Here, we analyzed endotracheal aspirate (ETA) scRNA-seq data (129K cells) from 40 intubated patients (both COVID-19 and non-COVID-19) in the COVID-19 Multiphenotyping for Effective Therapies (COMET) cohort. To understand the cell populations and gene programs involved in ARDS mortality in the lower airways, we performed both cell-type-agnostic and neutrophil-focused analyses using state-of-the-art computational methods, including CNA^36^ and cNMF.^37^ In the cell-type-agnostic analyses, we determined that airway neutrophils were a critical cell type prognostic for ARDS mortality. In the neutrophil-focused analyses, we identified a mortality-associated neutrophil subpopulation, in which up-regulated genes were involved in IFN-ɣ response (e.g., *TXNIP*, *IFI30*, *TRIM25*, *STAT1*, *RNF213*, and *TMEM140*), and ferroptosis (e.g., *PIM1*, *HMOX1*, *TXNRD1*, *GCLM*, and *FTL*), while down-regulated genes were involved in TNF-ɑ signaling (e.g., *PLAUR*, *IL1B*, *CCL20*, *GCH1*, *ICAM1*, *NFKBIA*, *TNFAIP6*, and *TNFAIP3*). We confirmed that this mortality-associated neutrophil subpopulation was associated with both COVID-19 and non-COVID-19 ARDS mortality, and was generally replicated in independent COVID-19 clinical data. Next, we determined that gene programs active in the mortality-associated neutrophil subpopulation could be recapitulated by cytokine stimulation *in vitro*. Finally, we confirmed that the mortality-associated neutrophil subpopulations could be induced in rodents in flu or SARS-CoV-2 infection models. Overall, our work provides additional insights into the role of neutrophils in ARDS.

## Results

### Cell-type agnostic analysis of 30-day mortality using the COMET ETA scRNA-seq data

We investigated the cell types involved in 30-day mortality (whether a patient died within 30 days of hospital admission), a pertinent ARDS clinical endpoint. To that end, we analyzed the endotracheal aspirate (ETA) scRNA-seq data collected from 40 (N_fatality_=10, N_survivor_=30) intubated COVID-19 and non-COVID-19 ARDS patients (see Table 1, Supplementary Table 1, Supplementary Figure 1). We applied Co-varying Neighborhood Analysis^36^ (CNA) to analyze the baseline samples (i.e., earliest samples taken since intubation), which consisted of 128,822 cells across 5 broad cell types. On average, these ETA samples were collected 2.1 days (s.d. 2.1) post ICU admission; CNA is a computational method to detect disease-associated cell states/populations at cell neighborhood resolution, and is more powerful than cell-type/cell-cluster based approaches.^36^ We elected to analyze the baseline samples to enrich for potential targetable biology and biomarkers for ARDS; later samples were more susceptible to the consequential effects of the disease and clinical interventions. We adjusted for known ARDS risk factors, e.g., age, sex, COVID-19 status, as well as other technical covariates, e.g., batch, time of sample collection in our analyses (see Methods). We elected not to adjust for corticosteroid use in our primary analyses, as this information was missing for 4 patients; and all but 1 of the remaining 36 patients received corticosteroid (Table 1**a**).

**Table 1:** Overview of the COMET ETA scRNA-seq data. (**a**) We report the number and proportion of the patients across demographic stratifications (e.g., age, sex, etc.), clinical outcomes (e.g., 30-day mortality status, max ordinal scale score), and treatment status (e.g., steroids and Tocilizumab). (**b**, **c**) We report the number and proportion of the major COMET ETA scRNA-seq cell types for all and non-COVID-19 patients, respectively. *Primary diagnoses of non-COVID-19 patients can be found in Supplementary Table 1.

| Patient characteristics | Number (proportion %) |  |  |
| --- | --- | --- | --- |
|  | All patients | non-COVID-19* | COVID-19 |
| **Sex at birth** |  |  |  |
| Male | 28 (70.0) | 7 (58.3) | 21 (75.0) |
| Female | 12 (30.0) | 5 (41.7) | 7 (25.0) |
| **Age (years)** |  |  |  |
| 18 – 64 | 32 (80.0) | 10 (83.0) | 22 (78.6) |
| 65 – 84 | 8 (20.0) | 2 (17.0) | 6 (21.4) |
| **Ethnicity** |  |  |  |
| Hispanic / Latino | 17 (42.5) | 2 (16.0) | 15 (53.6) |
| Non-Hispanic / Latino | 23 (57.5) | 10 (84.0) | 13 (46.4) |
| **Race** |  |  |  |
| Asian | 4 (10.0) | 2 (17.0) | 2 (7.1) |
| Black / African American | 4 (10.0) | 0 (0.0) | 4 (14.3) |
| Native Hawaiian / Other Pacific Islander | 1 (2.5) | 0 (0.0) | 1 (3.6) |
| White | 9 (22.5) | 6 (50.0) | 3 (10.7) |
| Other / Multiple Races | 22 (55.0) | 4 (33.0) | 18 (64.3) |
| **30-day mortality status** |  |  |  |
| Deceased | 10 (25.0) | 3 (25.0) | 7 (75.0) |
| Survived | 30 (75.0) | 9 (75.0) | 21 (25.0) |
| **COVID-19 status** |  |  |  |
| Positive | 28 (70.0) | 0 (0.0) | 28 (100.0) |
| Negative | 12 (30.0) | 12 (100.0) | 0 (0.0) |
| **Max ordinal scale score** |  |  |  |
| 6 | 2 (5.0) | 1 (8.3) | 1 (3.6) |
| 7 | 28 (70.0) | 8 (66.7) | 20 (71.4) |
| 8 | 10 (25.0) | 3 (25.0) | 7 (25.0) |
| **Treated with steroids** |  |  |  |
| True | 35 (87.5) | 7 (58.4) | 28 (100.0) |
| False | 1 (2.5) | 1 (8.3) | 0 (0.0) |
| Unknown | 4 (10) | 4 (33.3) | 0 (0.0) |
| **Treated with Tocilizumab** |  |  |  |
| True | 13 (32.5) | 0.0 (0.0) | 13 (46.4) |
| False | 23 (57.5) | 8 (66.7) | 15 (53.6) |
| Unknown | 4 (10.0) | 4 (33.3) | 0 (0.0) |

| Cell type | Number (proportion %) |  |
| --- | --- | --- |
|  | All time point | Earliest time point |
| Epithelial/Ciliated | 9,251 (3.0) | 8,261 (6.1) |
| Lymphocyte | 24,230 (8.0) | 15,279 (11.3) |
| Monocyte/Macrophage/Dendritic Cell | 154,327 (50.7) | 64,574 (47.6) |
| Neutrophil | 113,285 (37.2) | 47,018 (34.6) |
| Red Blood Cell | 3,259 (1.1) | 619 (0.4) |
| Total | 304,352 (100.0) | 135,751 (100.0) |

(c) Cell-type representations in non-COVID-19 patients
| Cell type | Number (proportion %) |  |
| --- | --- | --- |
|  | All time point | Earliest time point |
| Epithelial/Ciliated | 4,128 (8.4) | 3,460 (11.4) |
| Lymphocyte | 4,083 (8.2) | 3,119 (10.3) |
| Monocyte/Macrophage/Dendritic Cell | 29,075 (58.8) | 17,517 (57.9) |
| Neutrophil | 11,898 (24.1) | 5,947 (19.6) |
| Red Blood Cell | 252 (0.5) | 245 (0.8) |
| Total | 49,436 (100.0) | 30,288 (100.0) |

**Table 2:** Results from gene set enrichment analysis of the top 100 genes with the highest spectra scores of each cNMF topic. We report the top 5 MSigDB Hallmark gene sets (FDR < 0.05) for each gene program, except for gene program 4, which yielded less than 5 significant results. We report the complete results in Supplementary Table 7.

| cNMF gene program | Top 5 MSigDB Hallmark gene sets |
| --- | --- |
| 1 | Reactive Oxygen Species Pathway, Oxidative Phosphorylation, TNF-alpha Signaling via NF-kB, IL-2/STAT5 Signaling, Apoptosis |
| 2 | TNF-alpha Signaling via NF-kB, Inflammatory Response, Interferon Gamma Response, Apical Junction, Allograft Rejection |
| 3 | TNF-alpha Signaling via NF-kB, Allograft Rejection, Complement, Myc Targets V1, Inflammatory Response |
| 4 | Estrogen Response Early, Estrogen Response Late |

The results are reported in Figure 1, Supplementary Figure 2–3. Overall, airway neutrophils and lymphocytes had significantly positive (p < 0.01) average CNA neighborhood correlations (Figure 1**b, d**), suggesting an enrichment of these cell types in 30-day mortality cases. In contrast, other cell types (e.g., monocytes and epithelial cells) had significantly negative (p < 0.01) average CNA neighborhood correlation, indicating a depletion of these cell types in 30-day mortality cases. We note that although Red Blood Cells (RBCs) also had significantly positive (p < 0.01) average CNA neighborhood correlation, the proportion of RBCs was low in the scRNA-seq data (Table 1**b**, **c**; Supplementary Figure 2). We did not observe statistically significant differences in immune cell type proportions across 30-day mortality cases vs. controls (Supplementary Figure 2a). Although we observed a significant differential RBC proportion, the representation of RBC in the scRNA-seq data was limited (Table 1**b, c**, Supplementary Figure 2a).

**Figure 1:**
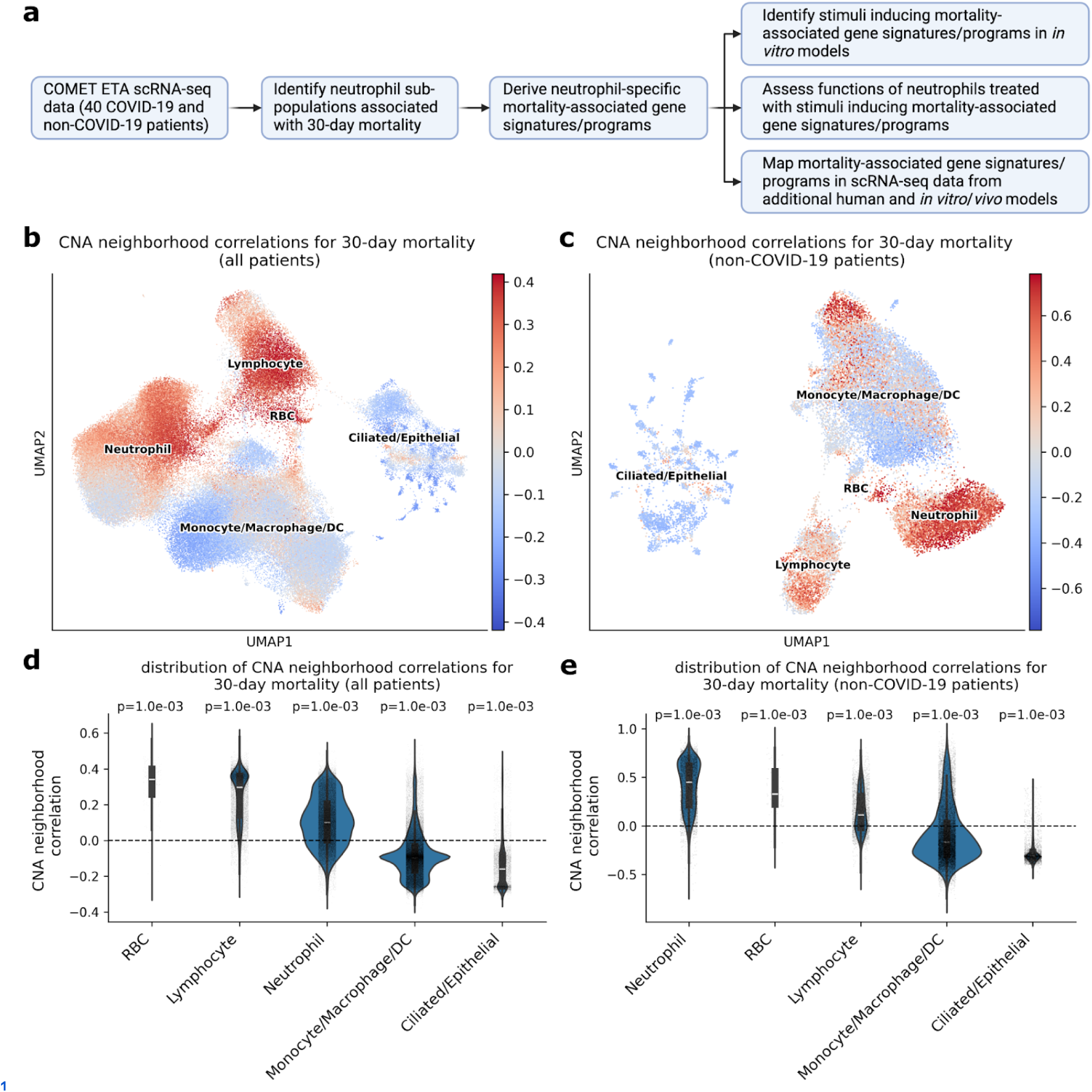
Results from cell-type agnostic analysis of 30-day mortality using the COMET ETA scRNA-seq data. (**a**) Flowchart showing the workflow of the overall study. (**b**, **c**) UMAP plots showing the CNA neighborhood correlations for 30-day mortality obtained using all and non-COVID-19 patients, respectively. (**d**, **e**) Violin plots showing the distribution of CNA neighborhood correlations across cell types in (**b**) and (**d**). respectively. The white lines in the violin plots represent the median, with boxes representing the interquartile range. The areas covered by the violins are proportional to the number of cells in each cell type. We report permutation p-values (based on 1,000 permutations), testing whether the average CNA neighborhood correlations of each cell type is significantly different from zero, on the top of each violin plot.

Next, we applied CNA specifically to 30,023 cells from the 12 non-COVID-19 patients (N_fatality_=3, N_survivor_=9), adjusting for age and sex, as well as technical covariates as above. Here, neutrophils had the highest average CNA neighborhood correlation (Figure 1**c, e**). Interestingly, the average lymphocyte CNA neighborhood correlation was much lower compared with the all-patient analysis (Figure 1**d**, **e**), while the results for other cell types were generally similar to the all-patient analysis (Figure 1**d, e**). We also did not observe statistically significant differences in cell type proportions across 30-day mortality cases vs. controls in the non-COVID-19 analysis (Supplementary Figure 2b).

We also performed a secondary analysis, in which corticosteroids use was included as an additional covariate to the ones adjusted in our primary analyses, using the 36 patients for whom this information is available. We observed similar disease-associated cell types and neutrophil subpopulations as those obtained without adjusting for corticosteroids use (Figure 1**b**, **d**, Supplementary Figure 3a, **b**). This is likely because the results were dominated by the patients (35 out of 36), who were treated with corticosteroids.

In summary, our CNA analysis of 30-day mortality using the COMET ETA scRNA-seq data suggested that neutrophils were strongly associated with severe ARDS outcomes in both COVID-19 and non-COVID-19 patients.

### ETA neutrophil-focused CNA analysis of 30-day mortality

Next, we performed neutrophil-focused analyses to identify neutrophil-specific ARDS disease biology, motivated by the strong association of ETA neutrophils with 30-day mortality in the cell-type agnostic analysis regardless of disease etiology (Figure 1**d**, **e**). We applied CNA to the 45,572 baseline ETA neutrophils across 37 (N_fatality_=8, N_survivor_=29) COVID-19 and non-COVID-19 patients post QC in the COMET scRNA-seq data (Table 1), adjusting for similar covariates as in the cell-type agnostic analyses (see above, Methods). We also identified neutrophil-specific mortality-associated genes that were correlated with the CNA neighborhood correlations across the neutrophils (see Methods). We elected not to perform lymphocyte-focused analysis, as the association with 30-day mortality was much weaker in the non-COVID-19 vs. all-patient analysis (Figure 1**d**, **e**); the relatively small number of lymphocytes (Table 1) also limited the statistical power of lymphocyte-focused analysis.

The results are reported in Figure 2, Supplementary Figure 4–11, and Supplementary Tables 2–6. The association with 30-day mortality among the neutrophils exhibited a gradient pattern, with a subset of neutrophils exhibiting higher CNA neighborhood correlation than others (Figure 2). As expected, the average CNA neighborhood correlation was higher for deceased vs. survived patients (Supplementary Figure 4).

**Figure 2:**
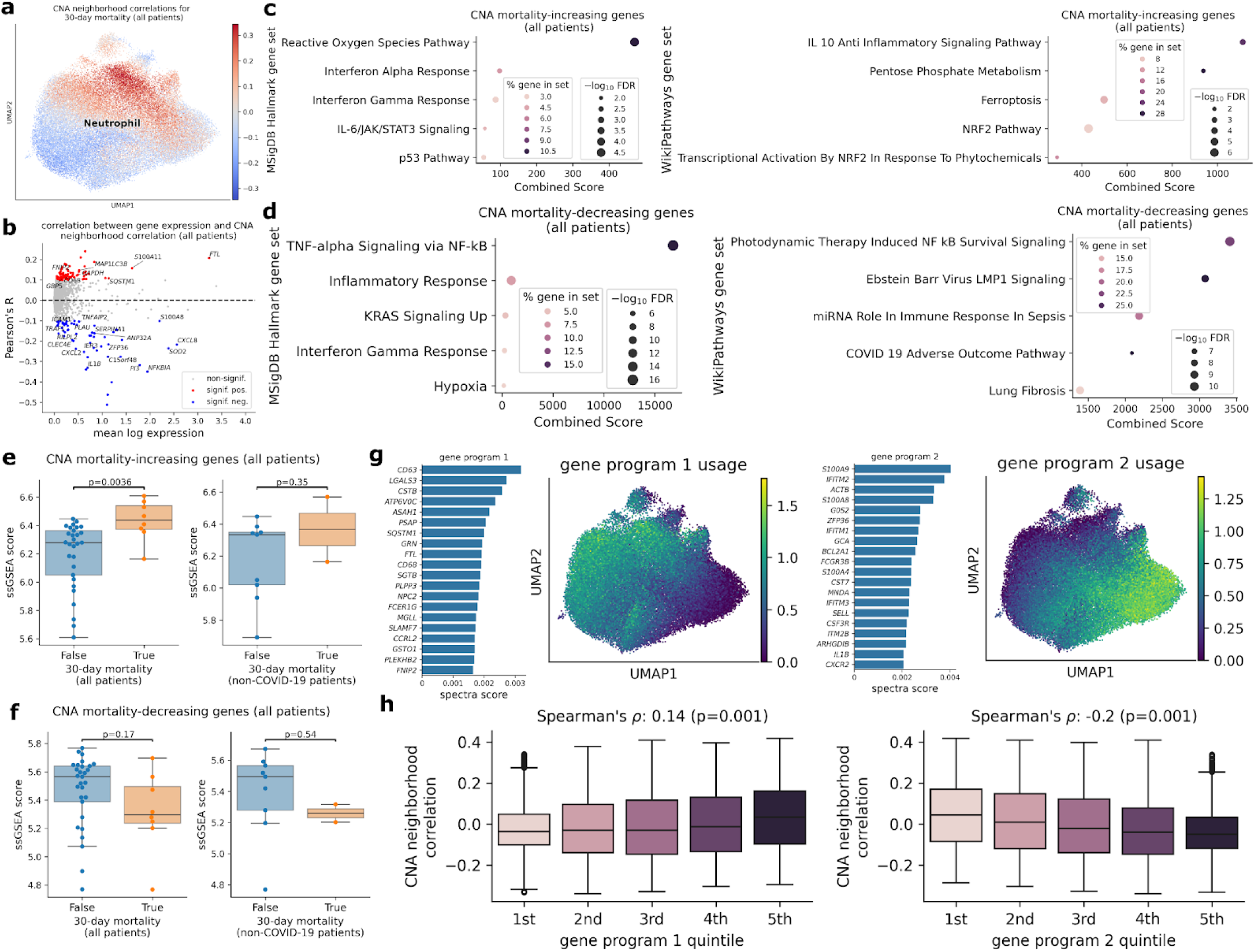
Results from the neutrophil-focused analysis of the COMET ETA scRNA-seq data using all patients. (**a**) We report the neutrophil-specific CNA neighborhood correlations for 30-day mortality obtained using all patients. (**b**) We report the correlation between the expression of each gene and CNA neighborhood correlations across the neutrophils. Genes with significant correlation (|R| > 0.1, FDR < 0.05) are highlighted as red and blue dots. (**c**, **d**) We report the top 5 MSigDB Hallmark (left) and WikiPathways (right) gene sets significantly (FDR < 0.05) overlapping genes positively and negatively correlated with the CNA neighborhood correlations, respectively. (**e**, **f**) We report the ssGSEA scores calculated using CNA mortality-increasing and decreasing genes, respectively, based on pseudobulked neutrophil expression from all patients (left panels) and non-COVID-19 patients (right panels). P-values were obtained using Wilcoxon rank-sum tests. (**g**) We report the top 20 genes with the highest spectra scores for gene program 1 (mortality-increasing genes) and program 2 (mortality-decreasing genes). The usage of the gene programs are shown in the UMAP plots. (**h**) We report the distribution of CNA neighborhood correlations in each quintile of the usages of gene program 1 and 2. The dark lines in the box plots represent the median, with the boxes representing the interquartile range (IQR), and whiskers representing 1.5× IQR.

However, many survived patients also harbored neutrophils with high CNA neighborhood correlation, and vice versa. This is expected, as CNA is designed to identify subtle neutrophil states associated with mortality, which could exist in both survived and decreased patients.

We identified 72 and 60 (out of 32,877) genes significantly positively (FDR < 0.05, R > 0.1) and negatively (FDR < 0.05, R < −0.1) correlated with the CNA neighborhood correlations across the cells (Supplementary Table 2), using similar approaches as introduced in ref.^36^. We refer to these genes as “CNA mortality-increasing genes” and “CNA mortality-decreasing genes” hereafter, respectively. Similar analyses using non-COVID-19 ARDS patients yielded smaller sets of mortality-associated genes that significantly overlapped those from the all-patient analysis (Supplementary Table 3, Supplementary Figure 5, <u>6</u>). Thus, we elected to use the larger gene sets derived from the better powered all-patient analysis for downstream analyses.

We performed several downstream analyses using the CNA mortality-associated genes. First, we performed over-representation analyses of the mortality-associated genes using GSEApy^38^, analyzing MSigDB Hallmark and WikiPathways gene sets. Both CNA mortality-increasing and mortality-decreasing genes significantly overlapped cytokine signaling gene sets, e.g., IFN-ɑ and IFN-ɣ response for the CNA mortality-increasing signature, and TNF-ɑ signaling via NF-κB for the CNA mortality-decreasing signature (Figure 2, Supplementary Table 4). We note the well-documented literature suggesting myeloid cells could be the cellular sources of IFN-ɣ^39,40^ and TNF-ɑ.^41,42^ Interestingly, the CNA mortality-increasing genes also significantly overlapped with reactive oxygen species and ferroptosis-related gene sets (Figure 2**c**, Supplementary Table 4**a**, **c**).

Second, we quantified the overall activity of the mortality-associated genes in each patient by applying ssGSEA^43,44^ to the pseudobulked neutrophil gene expression; higher ssGSEA scores indicate higher expression of the mortality-associated genes relative to other genes. As expected, ssGSEA scores for mortality-increasing genes were generally higher in 30-day mortality cases than controls, and vice versa for mortality-decreasing genes (Figure 2**e**, **f**). This was the case in both non-COVID-19 patients and in all patients (Figure 2**e**, **f**). We note that the lack of statistical significance in analysis of non-COVID-19 patients was likely attributable to a limited number of mortality cases.

Third, we investigated the cell types in which the CNA mortality-associated genes were active. We calculated AUCell^45^ scores for all the cells in the ETA scRNA-seq data using the CNA mortality-associated genes; higher AUCell scores represent higher gene activity. As expected, the activity of the CNA mortality-associated genes were specific to myeloid cells (Supplementary Figure 7). However, the activity of the CNA mortality-decreasing genes were generally specific to neutrophils, whereas the CNA mortality-increasing genes were also broadly active in monocytes, macrophages, and dendritic cells (Supplementary Figure 7). We also note that neutrophils with high AUCell scores for CNA mortality-increasing genes and high CNA neighborhood correlations did not fully overlap (Figure 2**a**, Supplementary Figure 7e, <u>8</u>). This was likely because genes characterizing neutrophils with high CNA neighborhood correlations were not captured by the gene signature, e.g., due to limited statistical power.

We also performed several additional secondary analyses to assess the robustness of the mortality-associated gene signatures. First, we calculated ssGSEA scores for each patient using mortality-associated genes derived from the remaining patients in a leave-one-out framework (Methods). The resulting ssGSEA scores for CNA mortality-increasing genes were generally higher in 30-day mortality cases than controls, and vice versa for mortality-decreasing genes (Supplementary Figure 9), consistent with the main results (Figure 2**f**). Second, we performed differential expression (DE) analysis contrasting 30-day mortality cases vs. controls via DESeq2,^46^ using the pseudobulked neutrophil gene expression (see Methods). As expected, the effects of the genes on mortality from pseudobulk DE analysis were generally consistent with those from CNA analysis (Supplementary Figure 10). Third, to identify proteins potentially driving the expression of the mortality-associated genes, we compared the ssGSEA scores for the CNA mortality-associated genes with the protein expression of IFN-ɣ and TNF-ɑ in ETA assayed using Olink^47^ (“Methods”). We note that IFN-ɑ, which was also implicated in our over-representation analysis (Figure 2**c**), is not included in the Olink protein assay panel. We observed weakly positive correlations between the ssGSEA scores and protein expression, with statistically significant result observed for the correlation between mortality-decreasing genes and TNF-ɑ (Supplementary Figure 11, Supplementary Table 6).

In summary, we identified a robust ETA neutrophil subpopulation associated with 30-day mortality in both COVID-19 and non-COVID-19 patients, characterized by up-regulation of genes involved in IFN-ɑ, IFN-ɣ response pathways, and ferroptosis, and down-regulation of genes involved in TNF-ɑ response pathway.

### Gene program analysis of ETA neutrophils

We investigated whether specific neutrophil functions and cellular processes were associated with 30-day mortality using consensus Nonnegative Matrix Factorization (cNMF).^37^ Briefly, cNMF decomposes the gene expression matrix from a single-cell study into gene programs, a set of genes that collectively contribute to a specific biological function or cellular process, and inferring their overall activity (i.e., usage of gene programs) in each individual cell. Unlike CNA, cNMF identifies gene programs in an unsupervised fashion, without explicitly modeling the disease outcome. We applied cNMF to the 45,572 neutrophils from all 37 patients (baseline samples) in the COMET ETA scRNA-seq data using cNMF to extract consistent gene programs across COVID-19 and non-COVID-19 patients (“Methods”).

The results of the cNMF analyses are reported in Figure 2, Supplementary Figures 12–14, and Supplementary Tables 7–8. We set the cNMF k parameter, the pre-specified number of gene programs, to 4, based on the cNMF k selection plot, balancing between stability of the factorization and expression matrix reconstruction error (Supplementary Figure 12a). We determined that the median usages of gene programs 1 and 2 were significantly (p=0.001) positively and negatively, respectively, correlated with the median AUCell scores for the CNA mortality-increasing genes, across all and non-COVID-19 patients (Figure 2**g**, Supplementary Figure 14). Notably, gene programs 1 and 2 were also the gene programs with the highest usages (Supplementary Figure 12b, **d**). Gene set enrichment analyses of the top 100 genes with the highest spectra scores for these gene programs revealed a set of shared pathways (e.g., IFN-ɣ response and TNF-ɑ signaling via NF-κB for both programs; Supplementary Table 8**a**, **b**, **e**, **f**), as well as a set of gene program specific pathways (e.g., ferroptosis and NRF2 pathways for gene program 1 only; Supplementary Table 8**e**). Despite the significant correlations, we note that the neutrophils with high CNA neighborhood correlations did not fully overlap those with high usage of gene program 1 and 2, suggesting that mortality-associated neutrophils likely involved multiple different cellular processes. We elected not to analyze gene programs 3 and 4 due to their relatively low usage across the ETA neutrophils (Supplementary Figure 12b, **d**).

In summary, our cNMF analysis of the ETA neutrophils suggested that ETA neutrophils expression could be captured by gene programs involving IFN-ɣ and TNF-ɑ signaling pathways, and that these gene programs partially explained the cell populations associated with 30-day mortality identified by CNA.

### Analysis of mortality-associated genes in independent COVID-19 clinical scRNA-seq data

We assessed the replicability of the CNA mortality-associated genes derived from the COMET ETA scRNA-seq data, in the publicly available COVID-19 scRNA-seq data from Bost et al.^48^ The Bost et al. data included 26,336 bronchoalveolar lavage fluid (BALF) and 21,652 blood neutrophils from 21 severe COVID-19 patients (N_fatility_=8, N_survivor_=12 for BALF; N_fatility_=7, N_survivor_=13 for blood; Supplementary Table 9). For each patient in the Bost et al. BALF or blood data, we calculated aggregated ssGSEA^43,44^ scores for CNA mortality-increasing and decreasing genes, based on the pseudobulked neutrophil gene expression.

The results for the analyses of the Bost et al. data are reported in Figure 3, Supplementary Figures 15–17, Supplementary Tables 10–11. In the analysis of the BALF neutrophils, the ssGSEA scores for CNA mortality-increasing were significantly positively associated with the SOFA scores (Spearman’s ⍴=0.49 (p=0.029), Figure 3**a**), and remained significant after adjusting for age and sex (Supplementary Table 10**a**). The association with mortality was also positive, but not statistically significant (p=0.067, Supplementary Figure 17a). We did not detect significant association between the ssGSEA scores for CNA mortality-decreasing genes and SOFA scores or mortality (Figure 3**b**, Supplementary Figure 16b, <u>17</u>**b**, **f**). In the analysis of the blood neutrophils, we did not detect a significant association between the ssGSEA scores for CNA mortality-associated genes and SOFA scores or mortality (Figure 3**c**, **d**, Supplementary Figure 16c, **d**, <u>17</u>**c**, **d**, **g**, **h**).

**Figure 3:**
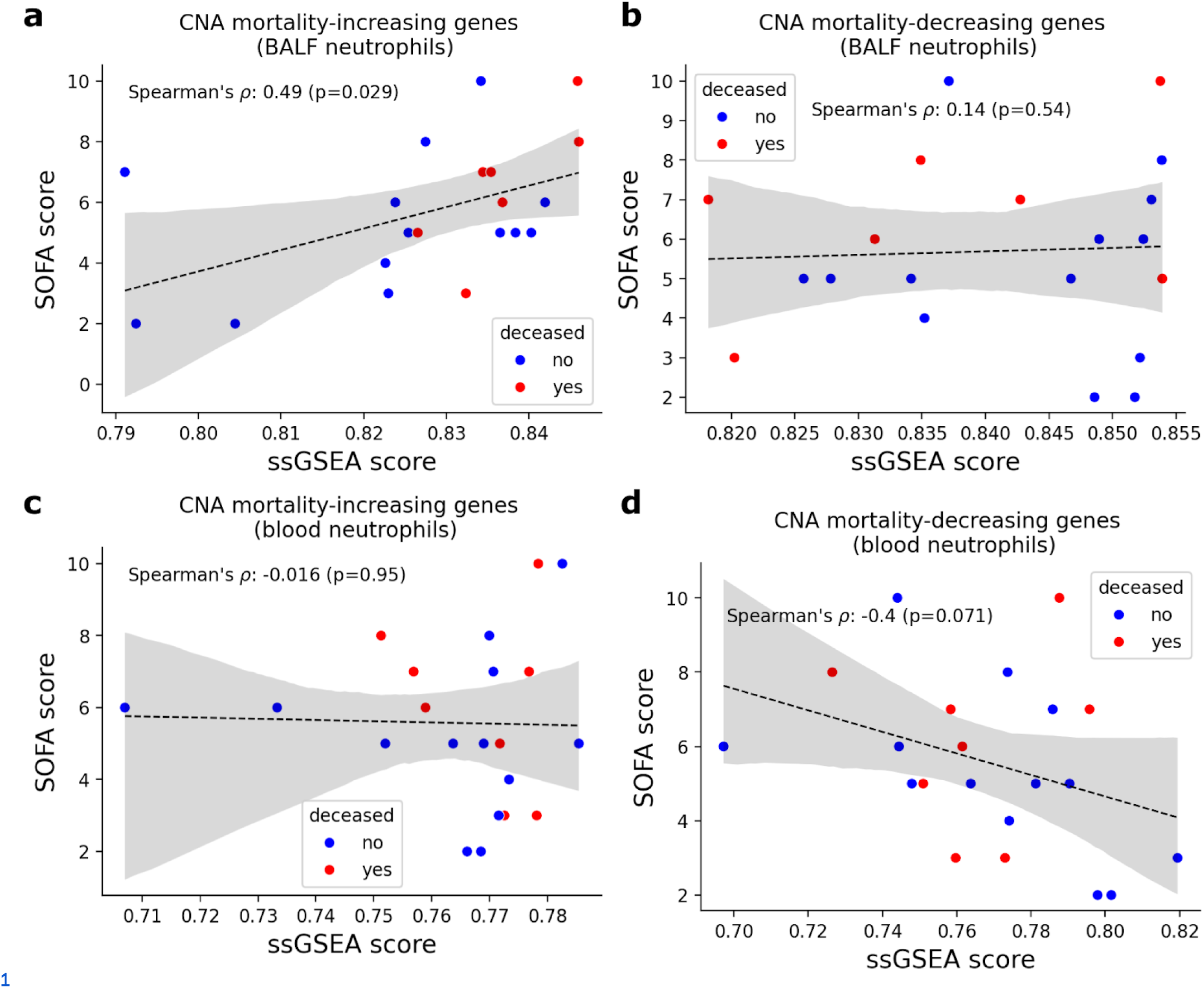
Results from the analysis of CNA mortality-associated genes in the neutrophils from the Bost et al. COVID-19 patient scRNA-seq data. (**a**, **b**) We report the Spearman’s correlation between patient SOFA clinical severity scores and ssGSEA scores for CNA mortality-increasing and decreasing genes, respectively, for BALF; we report analogous results for blood neutrophils in (**c**, **d**). ssGSEA scores were calculated based on log-normalized pseudobulk gene expression. Solid blue lines in the scatter plots are regression lines fitting the SOFA scores against the ssGSEA scores; shaded regions represent 95% confidence intervals on both sides.

As a secondary analysis, we also analyzed the CNA mortality-associated genes in a smaller publicly available COVID-19 scRNA-seq data from Liao et al.,^34^ which included 3,628 BALF neutrophils from 9 patients (Supplementary Table 12, Supplementary Figure 18). We calculated aggregated scores for CNA mortality-increasing and decreasing genes for each cell using AUCell.^45^ The aggregated CNA mortality-increasing and decreasing gene signature scores were both significantly higher in severe vs. mild patients (Supplementary Figure 19b, **d**).

In summary, the CNA mortality-increasing gene signatures derived from the analysis of 30-day mortality using the COMET ETA neutrophil scRNA-seq data were generally replicated in BALF, but not blood, neutrophils from independent COVID-19 scRNA-seq data. However, replication of the CNA mortality-decreasing gene signatures were generally weaker. We discuss ramifications of these results in Discussion.

### Analysis of mortality-associated genes using *in vitro* models

We next assessed the effects of TNF-ɑ, IFN-ɣ, dexamethasone, and lipopolysaccharide (LPS), on the expression of the CNA mortality-associated genes in human blood neutrophils in *in vitro* experiments; TNF-ɑ and IFN-ɣ are cytokines implicated in our pathway analyses, whose protein expression correlated with the expression of CNA mortality-associated genes in ETA (see subsection above); dexamethasone is a standard of care treatment for hospitalized COVID-19 patients and showed efficacy in reducing mortality;^13^ LPS is a pro-inflammatory stimulus for inducing sepsis in animal models.^49^

In the neutrophils treated with IFN-ɣ, 19 of the 72 CNA mortality-increasing genes were significantly (|log_2_ FC| > 0.5, FDR < 0.05) up-regulated and 13 down-regulated relative to medium (Figure 4**a**, **c**). The up-regulated CNA mortality-increasing genes exhibited significantly (p=0.03) stronger magnitude of log_2_ FC than down-regulated genes (Supplementary Figure 20a). In contrast, the magnitude of the log_2_ FC was similar for the 14 up-regulated and 18 down-regulated (out of 60) CNA mortality-decreasing genes (Supplementary Figure 20e)

**Figure 4:**
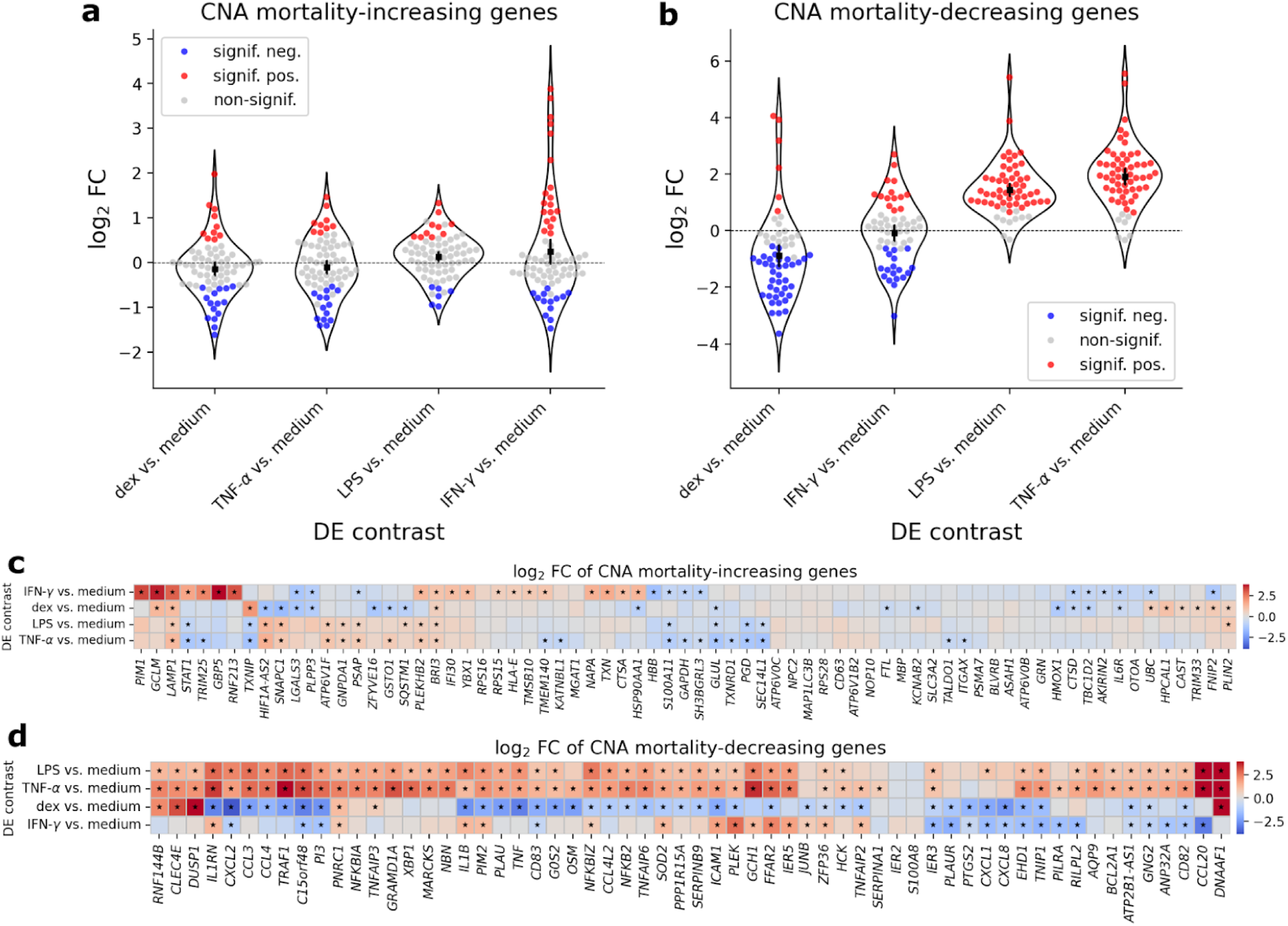
Results from the analysis of CNA mortality-associated genes in the *in vitro* blood neutrophil experiments. (**a, b**) We report the distributions of the log_2_ fold change (FC) of the expression of CNA mortality-increasing and decreasing genes relative to media, respectively, in neutrophils stimulated with dexamethasone (dex), TNF-ɑ, lipopolysaccharide (LPS), and IFN-ɣ. (**c**, **d**) We report the values of the log_2_ FC across the 4 contrasts, for the CNA mortality-increasing and decreasing genes, respectively. Red and blue dots in (**a**) and (**b**) represent significantly (|log_2_ FC| > 0.5, FDR < 0.05) up and down regulated genes, respectively; black dots and error bars in the center of the violin plots in (**a**, **b**) represent the average log_2_ FC and 95% confidence intervals (calculated using 1,000 bootstraped samples), respectively. Significant differentially expressed genes are marked by stars (“★”) in (**c**) and (**d**). Numerical results are reported in Supplementary Table 13.

In the neutrophils treated with TNF-ɑ, LPS, and dexamethasone, 9, 10, and 10 CNA mortality-increasing genes were up-regulated, respectively (Figure 4**a**, **c**). Importantly, these were generally distinct from those up-regulated by IFN-ɣ; 14, 6, and 15 genes were down-regulated, respectively (Figure 4**a**, **c**). We did not observe significant differences in the magnitude of log_2_ FC between up-regulated and down-regulated CNA mortality-increasing genes in the neutrophils treated with these stimuli(Supplementary Figure 20b, **c**, **d**).

Among the 60 CNA mortality-decreasing genes, 53 (0), 50 (0), and 6 (38) genes were up-regulated (down-regulated), respectively, in neutrophils treated with TNF-ɑ, LPS, and dexamethasone (Figure 4**b**, **d**, Supplementary Figure 20f, **g**, **h**).

We concluded that CNA mortality-increasing genes were likely induced by a combination of cytokines, whereas CNA mortality-decreasing genes could be induced by TNF-ɑ alone.

### Effects of IFN-ɣ and TNF-ɑ on neutrophil function and cytokine secretion

Motivated by the results from the analyses of transcriptomics data, we next performed *in vitro* experiments to assess the effects of IFN-ɣ, TNF-ɑ, and LPS on human neutrophils functions. We stimulated human blood neutrophils from 6 donors with TNF-ɑ, LPS, and IFN-ɣ, as well as combinations of IFN-ɣ with TNF-ɑ or LPS to mimic the complex cytokine environments in ARDS patients. We assessed cytokine release, and various immunological functions, including activation (indicated by reduced *CD62L* and *CD16*), degranulation (indicated by increased *CD63* and *CD66b*), and extracellular elastase activity.

The results are reported in Figure 5. We observed that IFN-ɣ alone had limited effect on neutrophil activation or degranulation (Figure 5**a**–**c**), while TNF-ɑ and LPS substantially induced both neutrophil functions (Figure 5**a**–**c**). However, when combined with either TNF-ɑ or LPS, IFN-ɣ enhanced neutrophil activation and degranulation (Figure 5**a**, **b**), indicating a synergistic effect. Observations of increased degranulation were also consistent with the elevated (although not statistically significant) activity of extracellular elastase, a byproduct of degranulation,^50,51^ in these conditions (Figure 5**d**).

**Figure 5:**
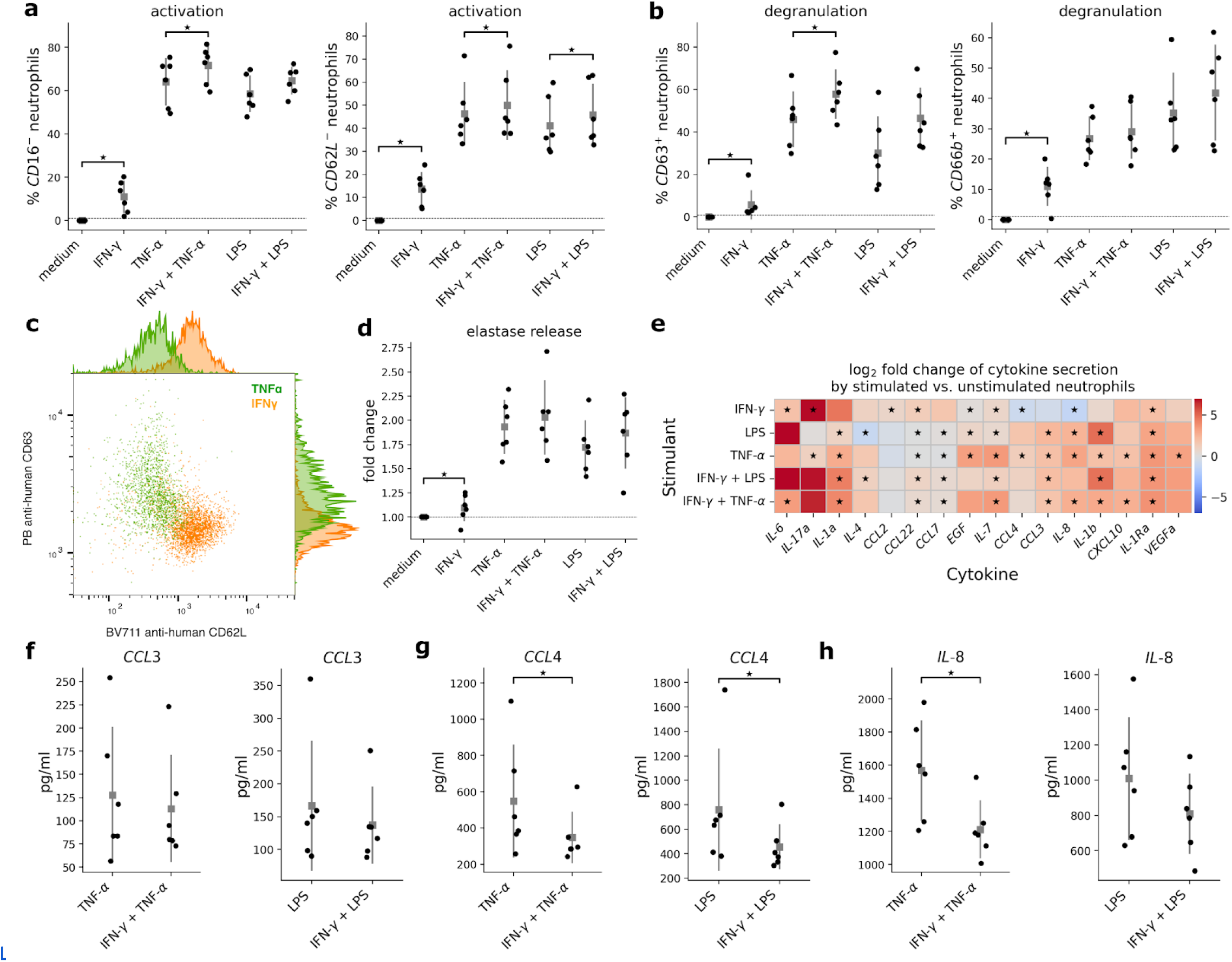
Functional and cytokine profiles of stimulated human primary blood neutrophils. (**a**) We report the proportions of neutrophils that were *CD16*^-^ (left) and *CD62L*^-^ (right), markers for neutrophil activation. (**b**) We report the proportions of neutrophils that were *CD63*^+^ (left) and *CD66b*^+^ (right), markers for neutrophil degranulation. (**c**) Flow cytometry plot showing neutrophil activation (*CD62L*^-^, x-axis) vs. degranulation (*CD63*^+^, y-axis,) for IFN-ɣ and TNF-ɑ stimulated neutrophils. (**d**) We report the fold change in measured elastase in stimulated vs. unstimulated (medium) neutrophils. (**e**) We report the log_2_ fold change in measured cytokine secretion by stimulated vs. unstimulated (medium) neutrophils. (**f**, **g**, **h**) We report the concentration (in picograms per milliliter) of *CCL3*, *CCL4*, and *IL-8*, respectively, in neutrophils stimulated with TNF-ɑ vs. IFN-ɣ + TNF-ɑ (left panels) and LPS vs. IFN-ɣ + LPS (right panels). Gray squares in the strip plots represent the average values; error bars represent one standard deviation on both sides. All p-values were obtained via the Wilcoxon signed-rank tests. Statistically significant results (p < 0.05) are denoted by “★”.

We also observed up-regulation of *CCL3*, *CCL4*, and *IL-8*, cytokines among the CNA mortality-decreasing genes, in neutrophils stimulated with TNF-ɑ or LPS, and down-regulation of these cytokines in neutrophils stimulated with IFN-ɣ (Figure 5**e**), consistent with our analysis of the bulk RNA-seq data (see subsection above). Notably, the expression of these cytokines were lower in neutrophils co-stimulated with IFN-ɣ and TNF-ɑ or LPS, compared to those stimulated with TNF-ɑ or LPS alone (Figure 5**f**–**h**), suggesting that IFN-ɣ could downregulate the production of these cytokines despite its synergistic effects on activation or degranulation. Finally, although IFN-ɣ had limited effect on activation, degranulation, or elastase release (Figure 5**a**–**c**), it up-regulated several cytokines, including *CCL2*, *CCL7*, *CCL22*, *IL-17a*, and *IL-1* (Figure 5**e**), indicating an impact on inflammatory signaling and further neutrophil recruitment.^52–55^

These results, along with our bulk RNA-seq results, suggest that IFN-ɣ and TNF-ɑ could synergistically influence neutrophil functions and gene programs, leading to a complex response that surpasses the effects of either cytokine alone.

### IFN-ɣ increased and TNF-ɑ decreased neutrophil ferroptosis

We investigated the role of IFN-ɣ and TNF-ɑ on neutrophil ferroptosis, a significant biological process implicated by pathway analysis of the CNA mortality-increasing signature (Supplementary Table 4). We analyzed the expression of both ferroptosis promoting genes, specifically, *LPCAT3*, *ACSL4*, *ALOX15*, *ALOX5*, *SAT1*,^56–61^ and ferroptosis inhibiting genes, specifically, *GPX4* and *GCLC*,^62^ in the bulk RNA-seq data for IFN-ɣ and TNF-ɑ treated neutrophils.

The results are reported in Figure 6 and Supplementary Table 13. IFN-ɣ significantly (FDR < 0.05) up-regulated ferroptosis promoting genes and down-regulated ferroptosis inhibiting genes, whereas TNF-ɑ had the opposite effect (Figure 6**a**). We note that the up/down regulation of ferroptosis promoting/inhibiting genes only entailed a ferroptosis-prone state, and did not confirm that the cells were actively undergoing ferroptosis.

**Figure 6:**
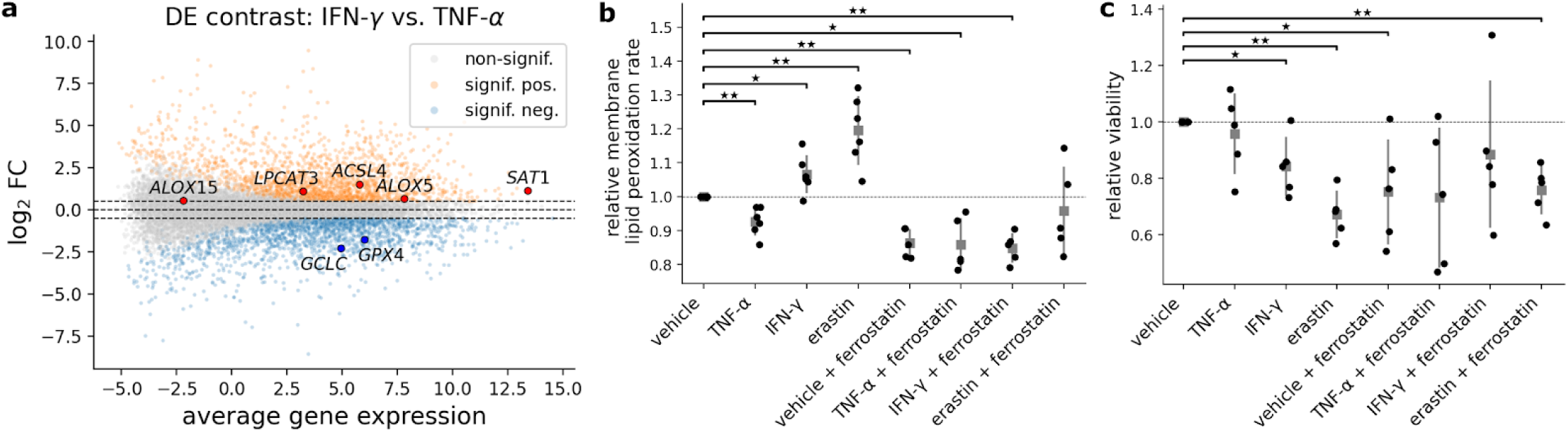
Transcriptional and functional profiles of ferroptosis in stimulated human primary blood neutrophils. (**a**) We report the results from DE analysis of bulk RNA-seq data of neutrophils stimulated with IFN-ɣ vs. TNF-ɑ. Significant DE genes (|log_2_ FC| > 0.5, FDR < 0.05) are highlighted in colors. Red and blue dots represent significant DE ferroptosis-inducing and inhibiting genes, respectively. (**b, c**) We report the membrane lipid peroxidization rates and relative viability (assayed via PI staining) of stimulated neutrophils. Each dot represents a donor. Gray squares in (**b**, **c**) represent the average values; error bars represent one standard deviation on both sides. Statistically significant differences are denoted by “★” (for p < 0.05) and “★★” (for p < 0.01).

To further investigate the role of IFN-ɣ and TNF-ɑ in ferroptosis, we measured oxidized lipids, a marker of ferroptosis, on the membrane of stimulated human primary neutrophils, across 6 biological replicates from different donors via flow cytometry. We observed that membrane lipid peroxidation was significantly increased (p<0.05) in IFN-ɣ stimulated neutrophils, and decreased (p<0.05) in TNF-ɑ stimulated neutrophils (Figure 6**b**). The membrane lipid peroxidation in IFN-ɣ stimulated neutrophils was reduced, after ferrostatin-1, a specific inhibitor of ferroptosis,^63^ was added (Figure 6**b**), confirming that the increase in membrane lipid peroxidation was indeed due to ferroptosis.

We also observed that IFN-ɣ significantly decreased neutrophil viability, whereas TNF-ɑ did not (Figure 6**c**). However, adding ferrostatin-1 in IFN-ɣ treated neutrophils did not completely prevent ferroptotic cell death (Figure 6**c**), suggesting that ferrostatin-1 was not sufficient to prevent cytotoxic effects of IFN-ɣ.

In summary, our results suggested that IFN-ɣ and TNF-ɑ could be potential inducers and inhibitors, respectively, of ferroptosis in human primary neutrophils.

### Analysis of mortality-associated genes using *in vivo* models

We investigated whether the mortality-associated genes derived from the human data could be recapitulated using *in vivo* models; being able to model the mortality-associated gene signatures *in vivo* can facilitate the development of therapeutics for ARDS. We analyzed publicly available BALF scRNA-seq data from Peidli et al.,^64^ which includes 12 hamsters infected with SARS-CoV-2 of varying doses (high vs. low) across 2 time points (3 biological replicates per (time point, dosage) combination; Supplementary Table 14). We also analyzed BALF and blood scRNA-seq data from a mouse influenza A (PR8) model, which includes 18 mice (3 replicate per condition, pooled into 1 sequencing batch) infected with 0, 10^3^, and 10^4^ plaque-forming unit (PFU) of flu virus, with samples collected at day 3 and day 7 (Supplementary Table 14, <u>15</u>, Supplementary Figure 21; Methods). For the Peidli et al. scRNA-seq data, we calculated ssGSEA^43,44^ scores for the CNA mortality-associated genes based on pseudobulked neutrophil gene expression of each hamster sample. For the mouse flu model scRNA-seq data, we calculated AUCell scores using the CNA mortality-associated genes for each individual cell; we were not able to calculate aggregated scores for each mouse sample as we did with the Peidli et al. data, as cells from multiple mice were pooled together during sequencing.

The results are reported in Figure 7 and Supplementary Figures 21–24. In the analysis of the Peidli et al. data, we observed significantly higher ssGSEA scores for CNA mortality-increasing genes in hamsters infected with high vs. low doses of SARS-CoV-2 at both day 2 and day 3 (Figure 7**a**); we did not observe significant changes in the ssGSEA scores for CNA mortality-decreasing genes across SARS-COV-2 doses at either time points (Figure 7**b**).

**Figure 7:**
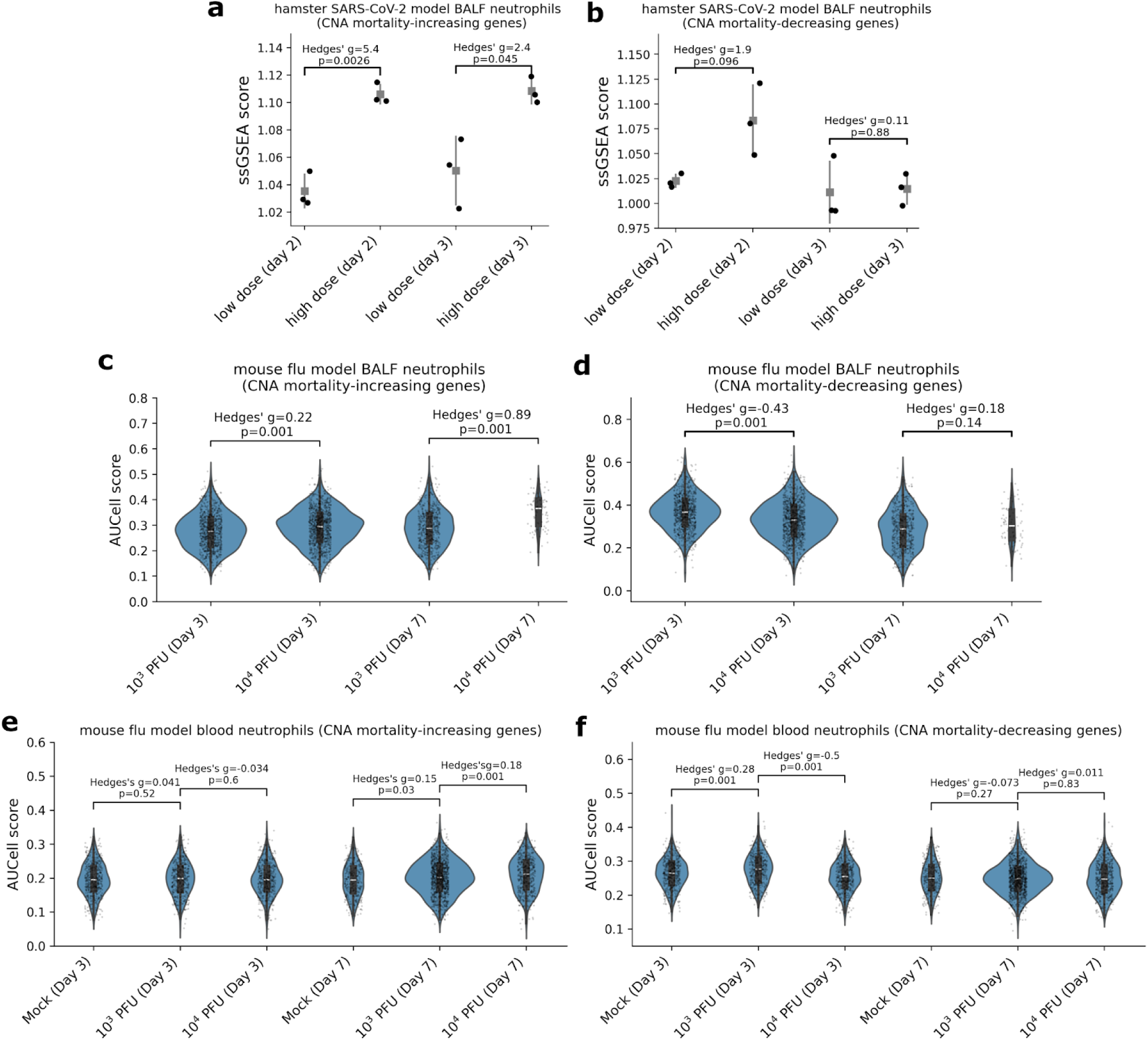
Results from the analysis of CNA mortality-associated genes in BALF and blood neutrophils from *in vivo* model scRNA-seq data. (**a**, **b**) We report the ssGSEA scores calculated using the CNA mortality-increasing and decreasing genes (mapped to mouse orthologs), respectively, across the 4 (dosage, timepoint) pairs, based on the pseudobulked BALF neutrophil gene expression, derived from the Peidli et al. hamster SARS-CoV-2 model scRNA-seq data. P-values were obtained via two-sided t-tests. Each dot represents a hamster replicate. Gray squares represent the average ssGSEA scores; error bars represent one standard deviation on both sides. (**c**, **d**) We report the AUCell scores calculated using the CNA mortality-increasing and decreasing genes, respectively, across 4 the (dosage, timepoint) pairs, based on the expression of each BALF neutrophil in the mouse flu model scRNA-seq data; analogous results obtained based on blood neutrophils are shown in (**e**, **f**). P-values were obtained via 1,000 permutations. Each dot represents a cell. The boxes within the violin plots represent the interquartile ranges; white solid lines in the boxes represent the median.

In the analysis of BALF neutrophils from our mouse influenza A model (Supplementary Figure 22), we observed that the AUCell scores for CNA mortality-increasing genes significantly increased with viral doses (p < 0.001) at both time points (Figure 7**c**, Supplementary Figure 23a, <u>24</u>**a**), with stronger effect at day 7 (Hedges’ g = 0.89) than day 3 (Hedges’ g = 0.22). We also observed significantly lower AUCell scores for CNA mortality-decreasing genes (p < 0.001) at day 3 but not at day 7 (p > 0.1) (Figure 7**d**, Supplementary Figure 23b, <u>24</u>**b**). We elected to exclude non-infected samples in our analyses of BALF data, as they yielded less than 50 neutrophils. In contrast, in the analysis of CNA mortality-increasing genes in blood neutrophils, we observed much weaker effect sizes in samples infected with high vs. low flu dose (Hedges’ g = 0.18 and −0.034 at day 7 and day 3, respectively; Figure 7**e**, Supplementary Figure 23c, <u>24</u>**c**). These results suggest that the association between mortality and CNA mortality-increasing genes may manifest in a tissue-specific manner (also see “Discussion”). Interestingly, we also observed significant lower AUCell scores for CNA mortality-decreasing genes in mice infected with high flu dose (Figure 7**f**, Supplementary Figure 23d, <u>24</u>**d**).

In summary, the mortality-associated gene signatures derived from analysis of human data were generally recapitulated in *in vivo* models, which can be used in future studies to test therapeutic hypotheses.

## Discussion

In this work, we highlight airway neutrophils as a key cell type associated with 30-day ARDS mortality, and identify mortality-associated neutrophil gene programs and signatures in the lower airway using CNA and cNMF. Notably, the mortality-associated neutrophil gene signatures were shared in ARDS patients of diverse etiology and not specific to COVID-19, and were also observed in independent COVID-19 patient cohorts and in rodents infected with SARS-CoV-2 or influenza virus. Pathway analyses implicated IFN-ɣ response and ferroptosis for CNA mortality-increasing genes, and TNF-ɑ response for mortality-decreasing genes. In *in vitro* experiments and functional assays of primary human neutrophils, IFN-ɣ upregulated a subset of the CNA mortality-increasing and ferroptosis-inducing genes, increased membrane lipid peroxidation (a marker of ferroptosis), while downregulating ferroptosis-inhibiting genes and decreasing neutrophil viability. Conversely, TNF-ɑ upregulated a subset of the CNA mortality-decreasing genes, while downregulating ferroptosis-inducing genes. Our *in vitro* experiments also suggest that IFN-ɣ and TNF-ɑ may interact to enhance the production of several chemokines, including *CCL3*, *CCL4*, and *CXCL8,* which could amplify recruitment of inflammatory cells to the lung.

Our study has several implications. First, our work implicates airway neutrophil ferroptosis, which could be induced by IFN-ɣ, as a critical component of ARDS disease biology based on analyses of human, *in vitro,* and *in vivo* experimental data. We postulate that reactive oxygen species (ROS) and peroxidized lipids, markers of ferroptosis^65^ that are damaging to the lung epithelium and endothelium,^66,67^ contribute to ARDS disease severity. We note that ferroptosis could also induce neutrophil recruitment.^67^ The increased abundance of neutrophils could subsequently elevate neutrophil extracellular traps (NETs) and proteases,^66,67^ both damaging to lung tissues,^28,68,69^ leading to more severe ARDS. Indeed, several cytokines involved in the recruitment of neutrophils, including *IL-17*,^70^ *IL-1ɑ*,^71^ *CCL3*,^72^ and *CXCL8*,^73^ were upregulated in IFN-ɣ stimulated neutrophils. We note, however, that additional studies (e.g., using *in vitro* and *in vivo* models) are necessary to assess the relative impact of different neutrophil biologies on ARDS severity. Second, our work also creates avenues for testing ARDS therapeutic strategies; we showed that the ARDS mortality-associated airway neutrophil phenotype in human data could be induced *in vitro* via IFN-ɣ stimulation and in rodent models *in vivo* via flu/SARS-CoV-2 infection. Third, our analyses suggest that the mortality-associated neutrophil phenotype is likely specific to the lung: an association between CNA mortality-increasing gene signature and disease severity/influenza infection status was only detected in BALF and not blood, in both the analysis of the Bost et al. human data and the analysis of the mouse flu model data. Fourth, our study suggests that complex cytokine interactions could be involved in ARDS severity/mortality. Specific neutrophil functions (e.g., activation and degranulation) were much stronger when neutrophils were co-stimulated with IFN-ɣ and TNF-ɑ vs. stimulated with IFN-ɣ alone; distinct cytokine secretion profiles were also observed when neutrophils were stimulated with IFN-ɣ and TNF-ɑ together vs. IFN-ɣ alone. Future work investigating the synergistic effects of cytokines on ARDS is warranted.

Finally, we discuss several limitations and caveats of this study. First, our all-patient analyses may suffer from reduced statistical power to detect mortality-associated disease biology, due to heterogeneity across COVID-19 and non-COVID-19 ARDS. Indeed, we observed different prioritizations of cell types (specifically between neutrophils and lymphocytes) in the all-patient vs. non-COVID-19 only analyses (Figure 1**d**, **e**). We note that our choice to analyze all patients was in part driven by the limited number of patients, and that the neutrophil-specific mortality-associated disease biology was similar across COVID-19 and non-COVID-19 ARDS. Second, the relatively smaller number of non-COVID-19 vs. COVID-19 ARDS in our data could bias the identification of the mortality-associated genes and disease biology towards COVID-19 ARDS. We mitigated this imbalance by regressing out COVID-19 status from 30-day mortality in our all-patient analyses. However, unadjusted residual heterogeneity in ARDS disease biology for COVID-19 vs. non-COVID-19 patients may still bias our results. Third, we did not account for race and ethnicity, which have been shown to be associated with ARDS severity,^74–76^ in our analyses, due to the limited number of sample sizes of specific minority populations in our study. Fourth, we also did not account for corticosteroids use, which has been shown to reduce ARDS mortality,^77,78^ in our analyses of the COMET ETA scRNA-seq data. However, since the majority of the patients in the COMET cohort were treated with corticosteroids, adjusting for this as a covariate had minimal impact on the cell types and cell subpopulations identified. Fifth, our pathway analyses of CNA mortality-decreasing genes implicated TNF-ɑ (Figure 2**d**, Supplementary Table 4), previously reported to promote ferroptosis,^67,79^ seemingly contradicting our conclusion that ferroptosis increases mortality. We note that the interaction between TNF-ɑ and IFN-ɣ more strongly induced ferroptosis than TNF-ɑ alone in our *in vitro* experiments. And it is possible that there was indeed stronger interaction between TNF-ɑ and IFN-ɣ, even though TNF-ɑ was comparably lower, in mortality cases vs. controls. Sixth, we note that while the effects of TNF-ɑ and IFN-ɣ on the CNA mortality-associated gene signatures were observed in our *in vitro* models, their effects remain uninvestigated in *in vivo* models, where the cytokine environments could be more complex. We also note that investigating the effects of other cytokines, e.g., IFN-ɑ and IL-6, etc., which were also implicated in our analysis (Figure 2**c**), may provide additional biological insights into the CNA mortality-associated genes. Seventh, we note that the CNA mortality-decreasing neutrophil gene signature derived from COMET ETA scRNA-seq data was significantly positively associated with COVID-19 severity in the analysis of BALF neutrophils from Liao et al.^34^ (Supplementary Figure 19d). We attribute this discrepancy to differences in the patient populations: relative expression of the gene signature could differ between severe vs. mild patients and mortality cases vs. controls among already severe patients. Eighth, we also note that replication was generally weaker for CNA mortality-decreasing vs. increasing gene signatures in the analysis of independent COVID-19 data (Figure 3) or *in vivo* model data (Figure 7). This suggests that mechanisms for survival/recovery could be more heterogeneous than those for mortality/deterioration, leading to overall weaker associations for the gene signature, although the sample sizes were also limited in these analyses. Ninth, we note that only a limited range of viral dosages were tested in the *in vivo* rodent models; and the resulting phenotype may better reflect ARDS severity instead of mortality. Tenth, we did not distinguish between subtypes of ARDS (e.g., hyperinflammatory vs. hypoinflammatory) in this work, other than COVID-19 vs. non-COVID-19 ARDS. Further reducing the heterogeneity in the ARDS patient populations could increase statistical power in detecting disease-associated biologies and lead to more effective treatments.^80–82^ Eleventh, we note that various forms of cell death (e.g., ferroptosis, pyroptosis, and necrosis) are interconnected and involve shared biological processes, e.g., production of ROS.^67^ Although our pathway analyses highlighted ferroptosis, other forms of cell death may also play critical roles in ARDS.^67^ Twelfth, we have not investigated the interactions between mortality-associated neutrophils with other cell types. Understanding the cross talk between these neutrophils and other cell types (e.g., epithelial and endothelial cells) may further facilitate the development of therapeutics for ARDS.^83,84^ Despite the limitations and caveats mentioned here, our study provides additional mechanistic insights into the role of airway neutrophils in ARDS mortality.

## Methods

### Overview of the COMET cohort

The COVID-19 Multi-Phenotyping for Effective Therapies (COMET) collected samples longitudinally from hospitalized patients presenting with COVID-19 symptoms, further described in ref.^25^. Clinical severity metrics include 30 day all-cause mortality and Sequential Organ Failure Assessment (SOFA) scores that were assessed at study enrollment (Day 0), when baseline samples were collected. The COMET study was approved by the Institutional Review Board: UCSF Human Research Protection Program (HRPP) IRB# 20-30497 and informed consent was obtained for all patients.

### COMET endotracheal aspirate (ETA) scRNA-seq data

The COMET ETA scRNA-seq data was collected as described in ref.^23^ (see Table 1 for the patient characteristics of the data). We started from the raw read counts from this dataset, removing all cells with fewer than 200 total reads or 200 detected genes. We also removed cells with percent mitochondrial reads greater than 20%. After the QC step, 304K cells remained. We then log-normalized the raw read count data using the scater package, and performed PCA using the 2,000 most variable genes. We corrected for batch effects using mnnCorrect,^85^ a batch effect correlation method based on mutual nearest neighbor (MNN). We performed shared nearest neighbor (SNN) based clustering using the 25 MNN dimensions; specifically, we used the `specFindClusters` function from Seurat v4.3.0.1,^86^ with the `R` parameter set to 2.0 and all other parameters set as the default. We manually annotated the cell types based on the expression of the cell type marker genes (Supplementary Figure 1). Prior to our main analyses, we subset for the 129K cells corresponding to the earliest sample for each patient.

### Identifying cell subpopulations associated with 30-day mortality

We performed co-varying neighborhood analysis^36^ (CNA) to identify cell subpopulations associated with 30-day mortality. Briefly, CNA determines that a cell neighborhood, defined as a group of cells with similar transcriptomics profile (e.g., within some random walks on the k-nearest-neighbor graph of the scRNA-seq data) is associated with severity, if the cell neighborhood is enriched for cells from more severe patients. Since CNA defines a cell neighborhood anchored at each cell, it enables the identification of severity-associated cell subpopulations at individual cell resolution.

In detail, we constructed k-nearest-neighborhood (k-NN) graphs for the cells in the scRNA-seq data based on the expression profiles of the top 2,000 highly variable genes (HVGs). We used the default settings of CNA to construct the neighborhood abundance matrices based on the k-NN graphs, and adjusted for appropriate covariates including age, sex at birth, COVID-19 test status, and batch. We did not adjust for corticosteroid use in our primary analyses, as this information was not available for 4 (out of 40) patients (Table 1**a**). However, we note that analyses with and without adjusting for corticosteroid use yielded similar results (see Results). We performed CNA at both all cell-type levels, identifying the most critical broad cell types for ARDS severity, and at neutrophil-specific level, identifying neutrophil subpopulations associated with ARDS. We also performed CNA analyses with all (both COVID-19 and non-COVID-1) patients included and analyses restricting to non-COVID-19 patients.

To identify genes indicative of a cell neighborhood’s association with mortality in neutrophils, we correlated the expression of each gene with the CNA neighborhood correlations across the cells. We defined CNA mortality-increasing and decreasing genes as the set of genes with R > 0.1 and R < 0.01, with two-tailed permutation based FDR < 0.05. We performed gene set enrichment analysis using the CNA mortality-associated genes using GSEApy^38^ to identify the biological processes enriched in mortality-associated cell populations.

### Leave-one-out CNA analysis

In the leave one-out framework, CNA was rerun as described above, but with one patient removed, and a CNA mortality-increasing signature and CNA mortality-decreasing signature derived through correlation of gene expression with the 30-day mortality CNA association statistic (FDR < 0.05, Pearson’s R > 0.1 for mortality-increasing, Pearson’s R < −0.1 for mortality-decreasing). The AUCell scores for each signature were then calculated for all patients. Once this process was repeated through all patients, the median leave-one-out AUCell score across all derived signatures was reported in Supplementary Figure 9.

### Inferring gene programs in ETA neutrophils

We inferred gene programs for ETA neutrophils using cNMF.^37^ Briefly, cNMF infers the sets of genes that collectively contribute to specific biological functions or cellular processes, and quantifies their overall activity (i.e., usages of the gene programs) in each individual cell based on non-negative factorization of the gene expression matrix. cNMF also requires a pre-specified number of factors, k, as the input.

We applied cNMF to the count data from our preprocessed ETA neutrophil scRNA-seq data (see subsection below), which included 45,572 neutrophils (baseline samples) from all 38 patients, using the top 2,000 highly variable genes to infer the non-negative factor. We determined the optimal value for k, by incrementally testing k from 2 to 10. For each k, we ran cNMF with 30 random starts with a maximum of 1,000 iterations per random start. We elected to set k to 4, as it yielded moderate reconstruction error of the gene expression matrix with high stability. We applied cNMF to all patients to infer a consistent set of gene programs across COVID-19 and non-COVID-19 patients. To glean biological insights into each gene program, we performed gene set overrepresentation analysis of the top 100 genes with the highest spectra scores of each gene program using GSEApy.^38^

### Differential gene expression analysis of 30-day mortality using pseudobulked ETA neutrophils

We performed differential expression analysis of 30-day mortality using pseudobulked ETA neutrophil gene expression. We restricted our analyses to the 31 patients with a minimum of 100 neutrophils in the samples of the earliest timepoint. For each patient, we obtained the pseudobulk expression as the sum of the raw read counts. We then used the PyDESeq2 package^87^ to perform normalization and differential expression analysis of the pseudobulked gene expression. In the analysis of 30-day mortality of all patients (N_fatality_=7, N_survivor_=24), we adjusted for age, sex, COVID-19 status, and timepoint as the covariates; in the analysis of non-COVID-19 patients (N_fatality_=2, N_survivor_=8), we adjusted for age, sex, and timepoint as covariates.

### Comparing IFN-γ and TNF-α protein expression vs. the expression of CNA mortality-associated genes

We obtained ETA protein expression data for a panel of 2,944 proteins from 17 patients using the Olink technology;^47^ 15 of these patients also had corresponding ETA scRNA-seq data (see Supplementary Table 6**a**). We obtained normalized protein expression (NPX) from the raw data using the approach as described in ref.^88^. We retained samples with more than 75% proteins above limit of detection (LOD) and with 2,000 proteins passing Olink’s default QC.

We calculated aggregated scores for CNA mortality-increasing and decreasing genes, respectively, based on the log-normalized pseudobulked ETA neutrophil gene expression, using ssGSEA^44^ for each of the 15 patients with both ETA protein expression and neutrophil scRNA-seq data. We compared the aggregated ssGSEA scores of CNA mortality-increasing and decreasing genes with the NPX of IFN-γ and IFN-α, respectively; IFN-γ and IFN-α signaling pathways were implicated in the gene set enrichment analysis of CNA mortality-increasing and decreasing genes, respectively (Results). We assessed the association between the ssGSEA scores and NPX both through Spearman’s rank correlation, and through linear models adjusting for age and sex.

### Mouse influenza A model lung and blood scRNA-seq data

We obtained lung and blood scRNA-seq data from influenza A (PR8) infected mice with varied viral doses (10^3^ and 10^4^ PFU) at day 3 and 7 post treatment; cells from 3 mice across both the 2 tissues and time points were pooled into the same pool using HTO multiplexing. A full table of HTO ids and corresponding samples is provided in Supplementary Table 15. All 6 blood sample pools were run through the same sequencing run, and all BALF samples were run together in a separate run. Reads were aligned and processed with the CellRanger v6.1.1 pipeline and bcl2fastq2-v2.20.0.422., with GRCm38 used as a reference and with ENSEMBL version 90 annotations.

Ambient RNA correction was applied to the data with CellBender (v0.2.0)^89^ with the following settings: `FPR` = 0.01, `epochs` = 80, `expected cells` = 37k, `expected droplets` = 52,000 for each sequencing pool. Following this, HTO demultiplexing was performed with HTODemux (Seurat v4.3.0.1); and droplets were removed if assigned as a doublet or if the difference between the expression score for the first and second assignment was less than 0.5 (i.e., an ambiguous droplet). Cells were then filtered so that each cell had at least 500 UMI counts and 400 detected genes, and with mitochondrial proportions below 3 median absolute deviation above median. scDblFinder (v1.14.0)^90^ was then used with the following parameters: `dbr.sd` = 1.0 (renders prior estimated doublet rate meaningless), `nfeatures` = 2500. scDblFinder was run separately for each sequencing run. Cell types were then labeled by marker gene expression (Supplementary Figure 21).

We then calculated AUCell^45^ scores for each neutrophils using the mouse orthologs of the CNA mortality-associated genes, and performed t-tests to compare tissue/dose/timepoint conditions, particularly between 10^3^ PFU and 10^4^ PFU for a fixed tissue and timepoint.

### *in vitro* human blood neutrophil bulk RNA-seq data

Neutrophils were isolated from peripheral blood of healthy donors (n=3) by negative selection via the StemCell Neutrophil Isolation Kit. These neutrophils were stimulated per 10^6^ cells with vehicle, TNF-ɑ (0.1μg/ml), LPS (0.1μg/ml), IFN-ɣ (0.1-1.0μg/ml) and combinations of IFN-ɣ with TNF-ɑ or LPS for 4 hours at 37 °C. All stimulations were performed in technical triplicates for each donor. RNA was isolated using the Qiagen RNeasy mini kit with additional DNase digestion. RNA was pooled from technical triplicate wells for each of 3 biological replicates (human donors). RNA purity and concentrations were analyzed via NaNoDrop. Bulk RNAseq was performed on all pooled samples. For each donor, RNA was isolated from 10^6^ freshly isolated neutrophils to compare changes in gene expression following stimulation, since pilot experiments show that gene expression and cell phenotypes are altered after culture at 37 °C.

RNA-seq data was processed as follows. First, reads with low nucleotide qualities (70% of bases with quality less than 23) or matches to rRNA and adapter sequences were removed using the HTSeqGenie software.^91^ Remaining sequencing reads were mapped to the reference human genome (GRCh38), using the GSNAP short read aligner.^92^ Expression counts per gene were quantified using HTSeqGenie,^91^ using ENSEMBL version 90 features.

### Neutrophil flow cytometry activation and degranulation analysis

Human peripheral neutrophils were isolated from healthy donors (n=6) as mentioned above. For each donor, 0.25*10^6^ neutrophils were stimulated with vehicles, TNF-ɑ (0.1μg/ml), LPS (0.1μg/ml), IFN-ɣ (0.1-1.0μg/ml) and combinations of IFN-ɣ with TNF-ɑ or LPS. After 4 hours of incubation, neutrophils were labeled with near-infrared live-dead dye (Invitrogen), SuperBright600 anti-human CD14, NovaFluorBlue 610-30s anti-human CD16, and BrilliantViolet711 anti-human CD62L, PacificBlue anti-human CD63. This antibody panel was verified with the EasyPanel software. Viable neutrophils were gated by subsequent gating for single cells (forward scatter area/forward scatter height), viable cells (live-dead staining). Activated neutrophils were expressed as a percentage of *CD62L*^-^ and *CD14*^-^ cells, degranulating neutrophils are expressed as percentage for *CD63*^+^ and *CD66b*^+^ cells.

### Luminex immunoassay for measuring cytokine release by stimulated neutrophils

A multiplexed Luminex immunoassay was used to map the cytokine release profile of the different neutrophil subpopulations. Human peripheral neutrophils were isolated from healthy donors (n=6) as mentioned above. For each donor, 2.5×10^5^ neutrophils were stimulated with vehicles, TNF-ɑ (0.1μg/ml), LPS (0.1μg/ml), IFN-ɣ (0.1-1.0μg/ml) and combinations of IFN-ɣ with TNF-ɑ or LPS. After 4 hours of incubation, supernatants were taken for cytokine measurement via Millipore Human 30-plex Luminex. Concentrations below detection limit were considered negative and were given a value one-half of the detection limit.

### Neutrophil elastase activity assay

Human peripheral neutrophils were isolated from healthy donors (n=6) as mentioned above. For each donor, 2.5×10^5^ neutrophils were stimulated with vehicles, TNF-ɑ, LPS, IFN-ɣ or combinations of these as described in the previous section. A fluorometric neutrophil elastase activity assay kit (Abcam) was used as instructed by the manufacturers. Duplicates and background controls were included for all samples in both assays. Values were normalized to vehicle conditions for each donor.

### Lipid peroxidation and viability assays to analyze ferroptosis in vitro

Human peripheral neutrophils were isolated from healthy donors (n=6) as mentioned above. For each donor, 2.5×10^5^neutrophils were stimulated with vehicles, TNF-ɑ (0.1 μg/ml) or IFN-ɣ (0.1μg/ml). Erastin (2μM), a known ferroptosis inducer, was added as positive control. Neutrophils were stimulated with each of these stimuli with and without the addition of ferrostatin-1 (1μM). Ferrostatin-1 is a synthetic anti-oxidant that inhibits ferroptosis in a two fold manner. It scavenging the alkoxyl radicals of oxidized lipids while oxidizing ferrous iron back to its less reactive form ferric iron. At the end of 4 hours incubation, lipid peroxidation was analysed through oxidation of BODIPY 581/591 C11 reagent via the Image-iT™ Lipid Peroxidation Kit (Thermofisher).

The ratio of lipid peroxidation was calculated as the ratio of oxidized lipids (MFI FITC) to both oxidized and reduced lipids (MFI Texas Red), analysed via flow cytometry. Values of lipid peroxidation were normalized to the vehicle conditions for each donor. Neutrophil viability was determined after 24h in culture with the above mentioned stimuli via PI staining. The number of viable cells was analysed via automatic counting chamber and the ratio of neutrophil viability was calculated after normalizing to vehicle conditions for each donor.

### Bost et al. COVID-19 bronchoalveolar lavage fluid (BALF) and blood scRNA-seq data

We obtained publicly available BALF and blood COVID-19 scRNA-seq data from 21 severe COVID-19 patients (N_fatility_=8, N_survivor_=12 for BALF; N_fatility_=7, N_survivor_=13 for blood; Supplementary Table 9) through Gene Expression Omnibus (GSE number: GSE157344) from ref.^48^. We performed log-normalization, PCA, and clustering^93^ on the raw read count data. We manually annotated the cells using lung cell type marker genes (Supplementary Figure 15). We then subset for the neutrophils, and obtained pseudobulked gene expression for patients with more than 25 neutrophils, yielding 20 patients for blood and BALF samples in total (19 overlapping patients between blood and BALF).

We performed ssGSEA scoring^44^ (gsva v1.50.0)^94^ on the pseudobulk gene expression data using the CNA mortality-increasing and decreasing genes, and assessed association between SOFA scores and ssGSEA scores through Spearman’s correlation (p-values obtained via permutation) and through modeling SOFA scores as a function of ssGSEA scores in linear models adjusting for age or age and sex (p-values obtained analytically). We also assessed association between the ssGSEA scores and death through comparing the mean ssGSEA scores of survived vs. deceased patients (p-values obtained via permutation) and through modeling mortality status as a function of the the ssGSEA scores in logistic regression models adjusting for age or for age and sex as covariates (p-values obtained analytically).

### Liao et al. COVID-19 BALF scRNA-seq data

We obtained publicly available BALF COVID-19 scRNA-seq data from 9 donors (5 severe, 3 mild, and 1 healthy control) through Gene Expression Omnibus (GSE number: GSE145926) from ref.^34^. We started from the raw read count data, filtering out cells that had fewer than 500 UMI counts or fewer than 200 detected genes, and setting upper thresholds for mitochondrial read fraction (above which droplets were excluded) at the minimum of 3 median absolute deviations or 0.25 (Supplementary Figure 18a–c). Once this initial filtering was done, we applied scDblFinder (v1.14.0) with the following parameters: `dbr`=NULL (i.e. doublet rate assumed to be 1% doublets for every 1,000 droplets in a sample), `dbr.sd`=0.0, `nfeatures`=2500, and batch set to sample ID (i.e., run scDblFinder fitting model for each batch separately) (Supplementary Figure 18d).

After the QC and doublet filtering, 65,187 singlets remained. We then log normalized the count data, ran PCA, corrected PCA coordinates via mutual nearest neighbors (MNN),^85^ and performed clustering based on a shared nearest graph in the MNN corrected space.^93^

We calculated AUCell scores (AUCell v 1.2.4.0)^45^ for a set of lung cell type markers (Supplementary Figure 18f), and examined the expression of *FCGR3B*. We annotated cluster 13 as neutrophils. We then calculated AUCell scores for each of the neutrophils 13 using CNA mortality-associated genes. We compared the mean AUCell scores across different severity, and obtained test statistics using permutation.

### Peidli et al. hamster SARS-CoV-9 model BALF scRNA-seq data

We obtained publicly available hamster SARS-CoV-9 model BALF scRNA-seq data from 12 hamsters across multiple infection dosages and time points through Gene Expression Omnibus (GSE number: GSE241133) from ref.^64^. We subset for neutrophils based on the cell type labels provided by the authors, and obtained pseudobulk neutrophil gene expression of the dataset, aggregating by tissue, dose, timepoint, and replicate (corresponding to one experimental run per set of aggregated cells).

We performed ssGSEA^44^ scoring for the CNA mortality-increasing and decreasing genes on the pseudobulk data using the gsva (v1.50.0) package.^94^ We compared the mean ssGSEA scores across different infection dosages for each timepoint, and obtained test statistics using two-sided t-tests.

## Data Availability

All data produced in the present study are available upon reasonable request to the authors

## Acknowledgements

The authors are grateful to K. Kiani, Y. Yang, W. Mu, and L. Orozco for helpful discussions.

## Author contributions

O.W., T.D., C.R., and H.S. designed the experiments. T.D. performed the experiments. O.W. and H.S. analyzed the data. O.W., T.D., and H.S. wrote the paper with assistance from all authors.

## Data and code availability

The Bost et al. COVID-19 bronchoalveolar lavage fluid (BALF) and blood scRNA-seq data is available on GEO with accession number GSE157344. The Liao et al. COVID-19 BALF scRNA-seq data is available on GEO with accession number GSE145926. data The Peidli et al. hamster SARS-CoV-9 infection model BALF scRNA-seq data is available on GEO with accession number GSE241133. Analysis scripts and additional experimental data generated in this study will be available upon reasonable request to the authors.

