## Supplementary Information for "Identification of robust mortality-associated neutrophil gene programs and cytokine responses using large-scale ARDS endotracheal aspirate scRNA-seq data"

### Supplementary Tables

Supplementary Table 1: **Additional information for the COMET ETA scRNA-seq data.** (**a**) We report the primary diagnoses for each of the 12 non-COVID-19 patients in the COMET cohort. (**b**) We report the distribution of the earliest sample collection timepoint (days since ICU admission) across the patients.

[Supplementary_Table_1.xlsx]

Supplementary Table 2: **Genes significantly correlated with CNA neighborhood correlation from analysis of neutrophils of all patients.** (**a**) Genes significantly (FDR < 0.05, R > 0.1) positively correlated with CNA neighborhood correlations. (**b**) Genes significantly (FDR < 0.05, R < -0.1) negatively correlated with CNA neighborhood correlations.

[Supplementary_Table_2.xlsx]

Supplementary Table 3: **Genes significantly correlated with CNA neighborhood correlation in analysis of neutrophils from non-COVID-19 patients.** (**a**) Genes significantly (FDR < 0.05, R > 0.1) positively correlated with CNA neighborhood correlations. (**b**) Genes significantly (FDR < 0.05, R < -0.1) negatively correlated with CNA neighborhood correlations.

[Supplementary_Table_3.xlsx]

Supplementary Table 4: **Results of gene set enrichment analysis of genes significantly correlated with CNA neighborhood correlations in analysis of neutrophils of all patients.** (**a, b**) MSigDB Hallmark gene sets significantly (FDR < 0.05) overlapping genes significantly positively and negatively correlated with CNA neighborhood correlations, respectively. (**c, d**) WikiPathway gene sets significantly (FDR < 0.05) overlapping genes significantly positively and negatively correlated with CNA neighborhood correlations, respectively.

[Supplementary_Table_4.xlsx]

Supplementary Table 5: **Results of gene set enrichment analysis of genes significantly correlated with CNA neighborhood correlations in analysis of neutrophils of non-COVID-19 patients.** (**a, b**) MSigDB Hallmark gene sets significantly (FDR < 0.05) overlapping genes significantly positively and negatively correlated with CNA neighborhood correlations, respectively. (**c, d**) WikiPathway gene sets significantly (FDR < 0.05) overlapping genes significantly positively and negatively correlated with CNA neighborhood correlations, respectively.

[Supplementary_Table_5.xlsx]

Supplementary Table 6: **Association between ssGSEA scores for CNA mortality-associated genes and the protein expression of IFN-ɣ and TNF-α.** (**a**) We report the patient metadata, the ssGSEA scores for CNA mortality-associated genes, and the protein expression (Olink NPX values) of IFN-ɣ and TNF-α. (**b**, **c**) We report the association between ssGSEA scores for CNA mortality-increasing genes and the protein expression of IFN-ɣ and TNF-α, respectively. (**d**, **e**) We report the association between ssGSEA scores for CNA mortality-decreasing genes and the protein expression of IFN-ɣ and TNF-α, respectively. We evaluated the association between the ssGSEA scores and protein expression under linear models, adjusting for age and sex as covariates.

[Supplementary_Table_6.xlsx]

Supplementary Table 7: **Top 100 genes with the highest cNMF spectra score for each inferred gene program in analysis of ETA neutrophil scRNA-seq data.** (**a**, **b**, **c**, **d**) We report the top 100 genes with the highest cNMF spectra scores for cNMF gene program 1, 2, 3, and 4, respectively.

[Supplementary_Table_7.xlsx]

Supplementary Table 8: **Results of gene set enrichment analysis of the top 100 genes with the highest cNMF spectra score for each inferred gene program (cNMF on neutrophils for all patients).** (**a**, **b**, **c**, **d**) MSigDB Hallmark gene sets significantly (FDR < 0.05) overlapping the top 100 genes with the highest cNMF spectra scores for gene program 1, 2, 3, and 4, respectively. (**e**, **f**, **g**, **h**) WikiPathway gene sets significantly (FDR < 0.05) overlapping the top 100 genes with the highest cNMF spectra scores for gene program 1, 2, 3, and 4, respectively.

[Supplementary_Table_8.xlsx]

Supplementary Table 9: **Metadata of the patients from the Bost et al. 2021 COVID-19 scRNA-seq data.** (**a**, **b**) We report the metadata for the patients from the Bost et al. 2021 COVID-19 blood and lung neutrophil scRNA-seq data, respectively. For each tissue, we excluded patients with fewer than 25 neutrophils.

[Supplementary_Table_9.xlsx]

Supplementary Table 10: **Association between ssGSEA scores for CNA mortality-associated genes and SOFA scores in the analysis of Bost et al. COVID-19 scRNA-seq data.** (**a**, **c**) We report the association between SOFA scores and ssGSEA scores for CNA mortality-increasing and decreasing genes, respectively, calculated based on pseudobulked lung neutrophils of all patients; we report analogous results for the blood neutrophils in (**e**, **g**). (**b**, **d**) We report the association between SOFA scores and ssGSEA scores for CNA mortality-increasing and decreasing genes, respectively, calculated based on pseudobulked lung neutrophils of male patients only; we report analogous results for the blood neutrophils in (**f**, **h**). We evaluated the association between the ssGSEA scores and SOFA scores under linear regression models, adjusting for both age and sex as covariates in (**a**, **c**, **e**, **g**), and adjusting for age only in (**b**, **d**, **f**, **h**).

[Supplementary_Table_10.xlsx]

Supplementary Table 11: **Association between ssGSEA scores for CNA mortality-associated genes and mortality status in the analysis of Bost et al. COVID-19 scRNA-seq data.** (**a**, **c**) We report the association between mortality status and ssGSEA scores for CNA mortality-increasing and decreasing genes, respectively, calculated based on pseudobulked lung neutrophils of all patients; we report analogous results for the blood neutrophils in (**e**, **g**). (**b**, **d**) We report the association between mortality status and ssGSEA scores for CNA mortality-increasing and decreasing genes, respectively, calculated based on pseudobulked lung neutrophils of male patients only; we report analogous results for the blood neutrophils in (**f**, **h**). We evaluated the association between the ssGSEA scores and mortality status under logistic regression models, adjusting for both age and sex as covariates in (**a**, **c**, **e**, **g**), and adjusting for age only in (**b**, **d**, **f**, **h**).

[Supplementary_Table_11.xlsx]

Supplementary Table 12: **Characteristics of the patients in Liao et al. COVID-19 scRNA-seq data.** We report the disease severity as well as the numbers of neutrophils for each patient in the Liao et al. COVID-19 scRNA-seq data.

[Supplementary_Table_12.xlsx]

Supplementary Table 13: **Results from the differential expression analysis of the *in vitro* model experiments.** We report the $\log_2$ fold change, mean expression, and test statistics from the differential expression analysis contrasting (**a**) neutrophils stimulated with dexamethasone (dex) vs. media, (**b**) neutrophils stimulated with IFN-ɣ vs. media, (**c**) neutrophils stimulated with lipopolysaccharide (LPS) vs. media, (**d**) neutrophils stimulated with TNF-α vs. media, (**e**) neutrophils stimulated IFN-ɣ vs. TNF-α.

[Supplementary_Table_13.xlsx]

Supplementary Table 14: **Number of neutrophils for each sample in the scRNA-seq data from *in vivo* models.** (**a, b**) We report the number of neutrophils for blood and lung samples, respectively, in the mouse flu model scRNA-seq data. (**c**) We report the number of neutrophils for lung samples in the Peidli et al. hamster COVID-19 model scRNA-seq data.

[Supplementary_Table_14.xlsx]

Supplementary Table 15: **HTO tags and sequencing pools for each sample in the mouse flu model experiment.** We report the mapping from sequencing pools/samples to HTO tag IDs and experimental conditions.

[Supplementary_Table_15.xlsx]

### Supplementary Figures

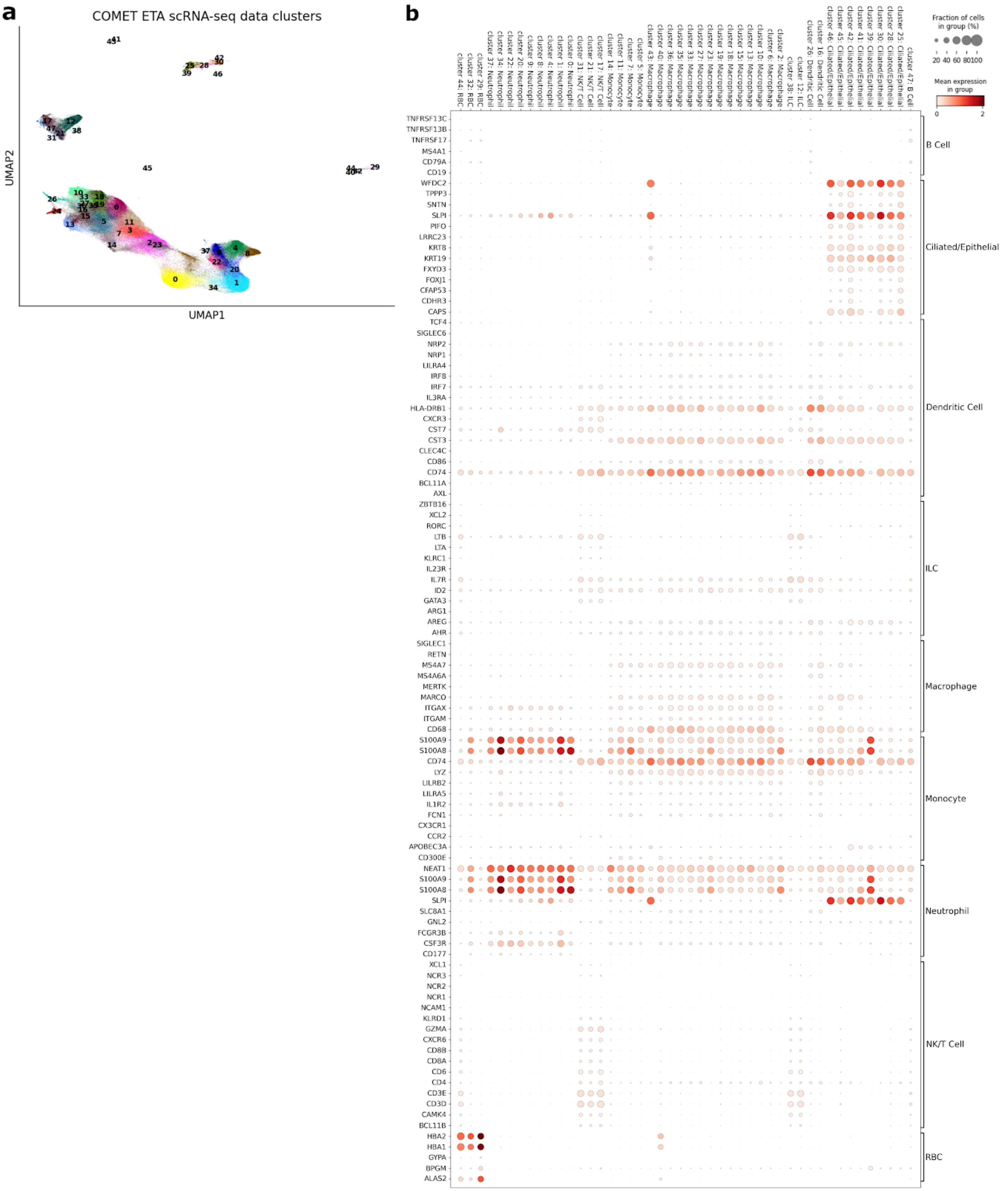

Supplementary Figure 1: **Annotating the COMET ETA scRNA-seq data.** (**a**) Clusters obtained for the COMET ETA scRNA-seq data (see Methods for the details of the clustering procedure). (**b**) Dot plot of mean expression and fraction of cells expressing for each (gene, cluster) pair, with clusters (and annotated cell types) indicated in columns, and genes indicated by rows. Groups of genes used as markers for particular cell types are indicated by brackets on the right-hand side.

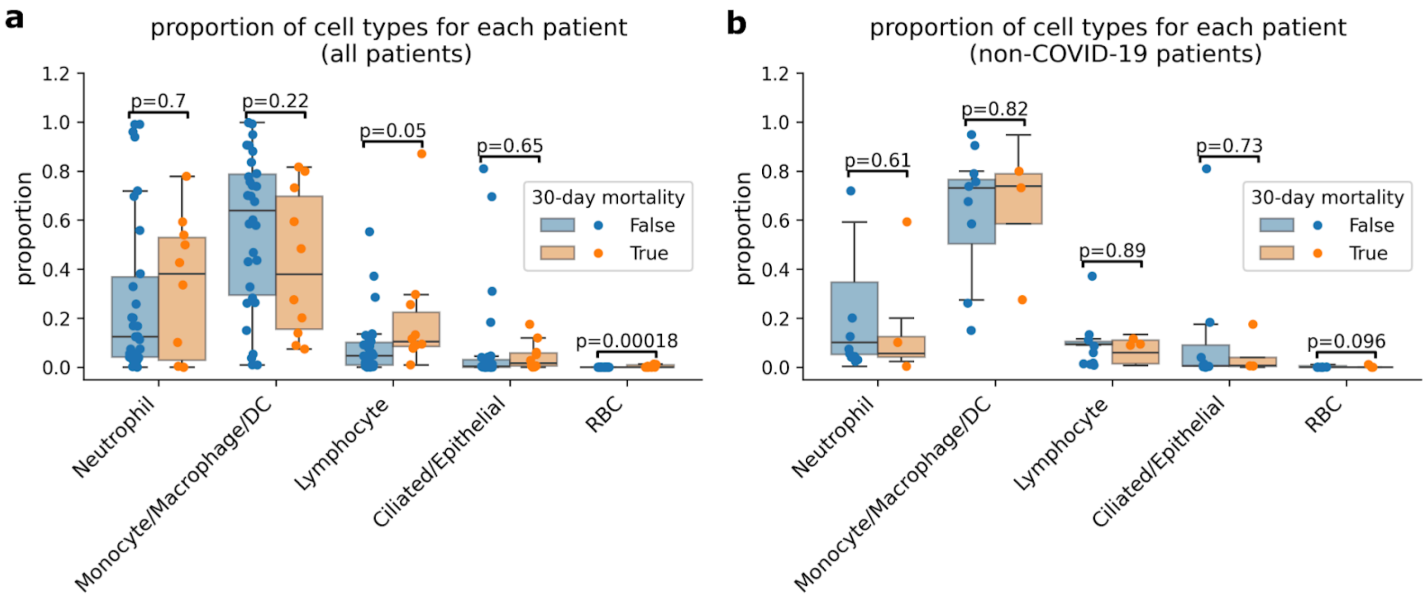


Supplementary Figure 2: **Proportion of each cell type from the patients in the COMET ETA scRNA-seq data.** (**a**) We report the proportions of cell types across all patients. (**b**) We report the proportions of cell types across non-COVID-19 patients. The dark lines in the box plots represent the median, with the boxes representing the interquartile range (IQR), and whiskers representing 1.5× IQR. P-values were obtained using Student's t-test with 1 degree of freedom.

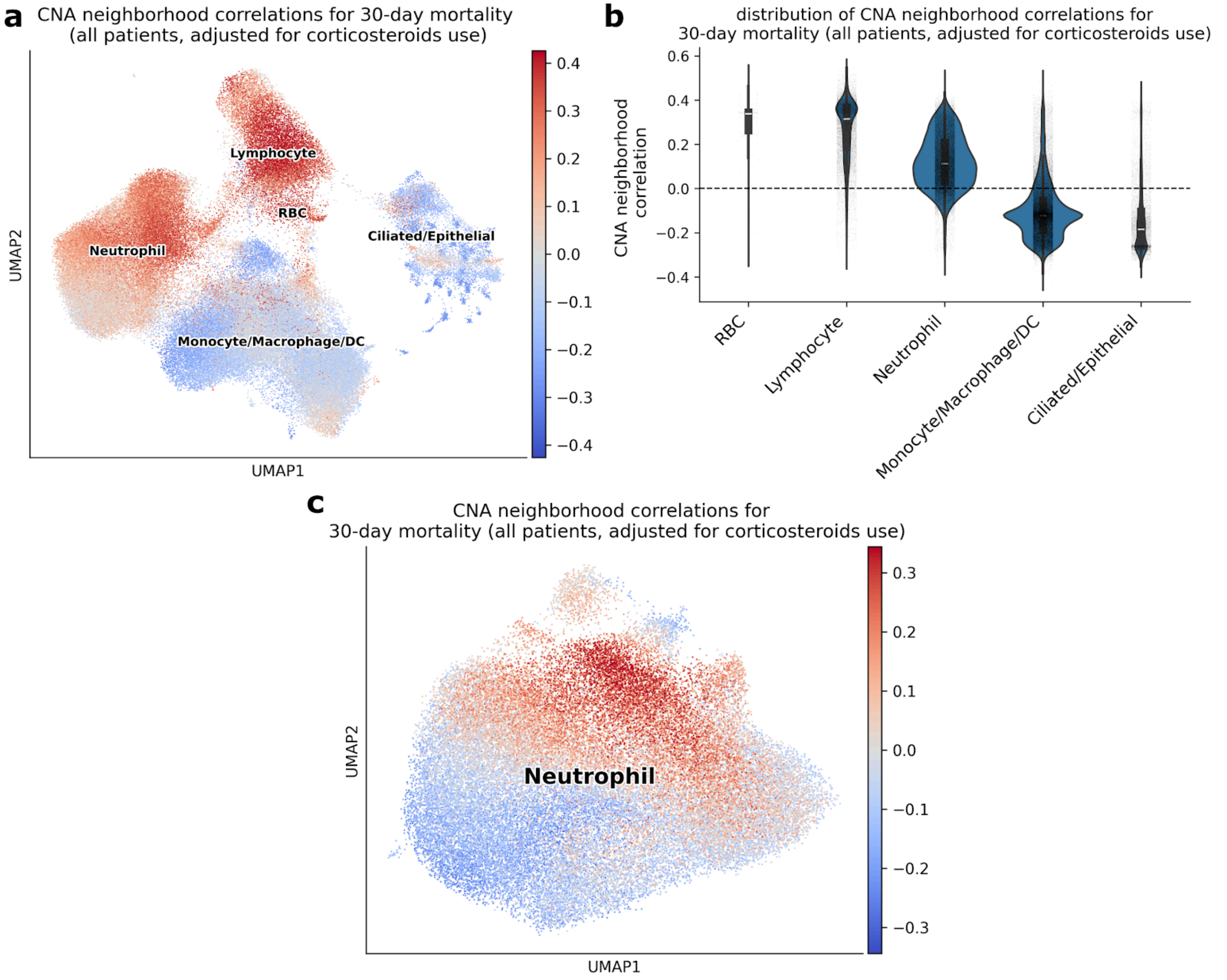


Supplementary Figure 3: **Results from analysis of 30-day mortality using COMET ETA scRNA-seq data adjusting for corticosteroids use.** (**a**) UMAP plots showing the CNA neighborhood correlations for 30-day mortality obtained using all patients in all COMET ETA cell types. (**b**) Violin plot showing the distribution of CNA neighborhood correlations across cell types in (**a**). The white lines in the violin plots represent the median. (**c**) MAP plots showing the CNA neighborhood correlations for 30-day mortality obtained using all patients in neutrophils.

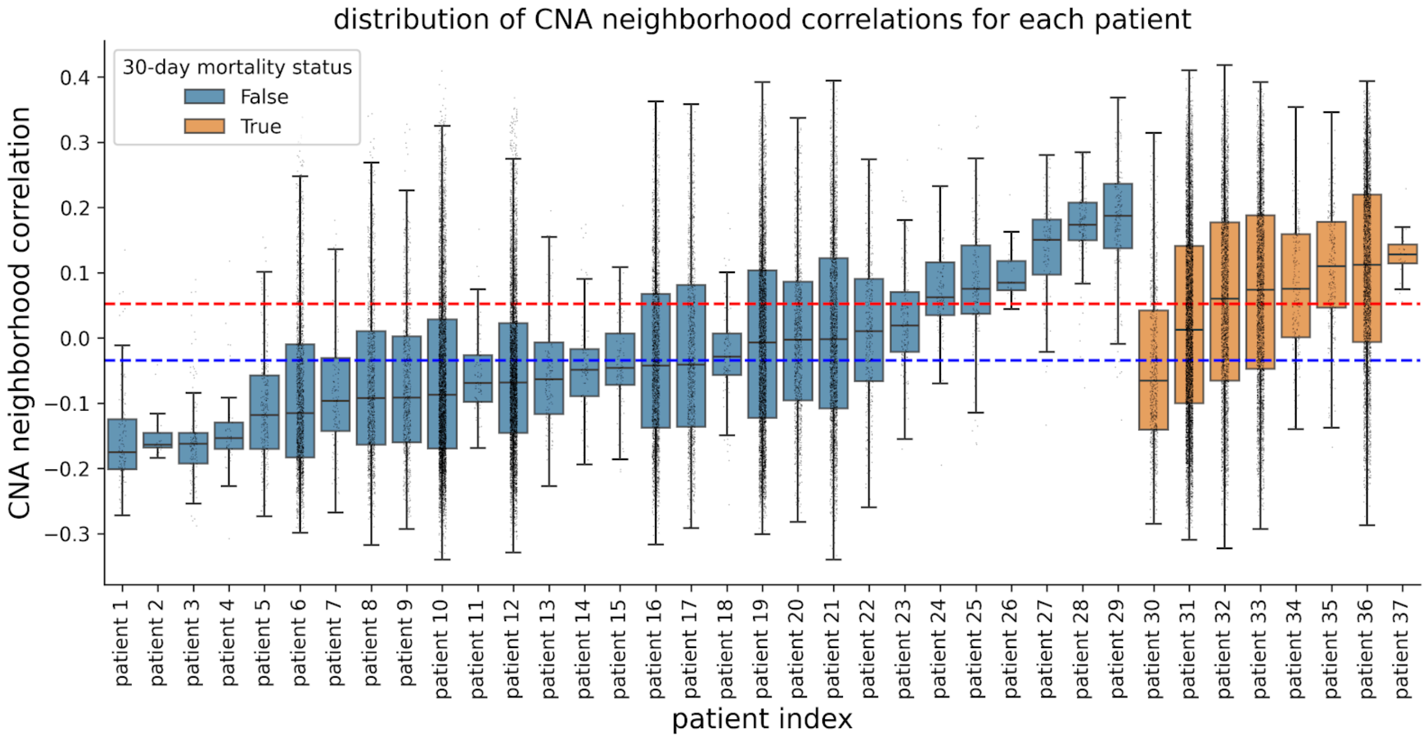


Supplementary Figure 4: **Distribution of COMET ETA neutrophil CNA neighborhood correlations for 30-day mortality.** Each dot in the box plot represents a neutrophil. The dark lines in the box plots represent the median, with the boxes representing the interquartile range (IQR), and whiskers representing 1.5× IQR. Donors are sorted based on the median CNA neighborhood correlation. Blue and red dashed lines represent the average CNA neighborhood correlation across all survived and deceased patients, respectively.

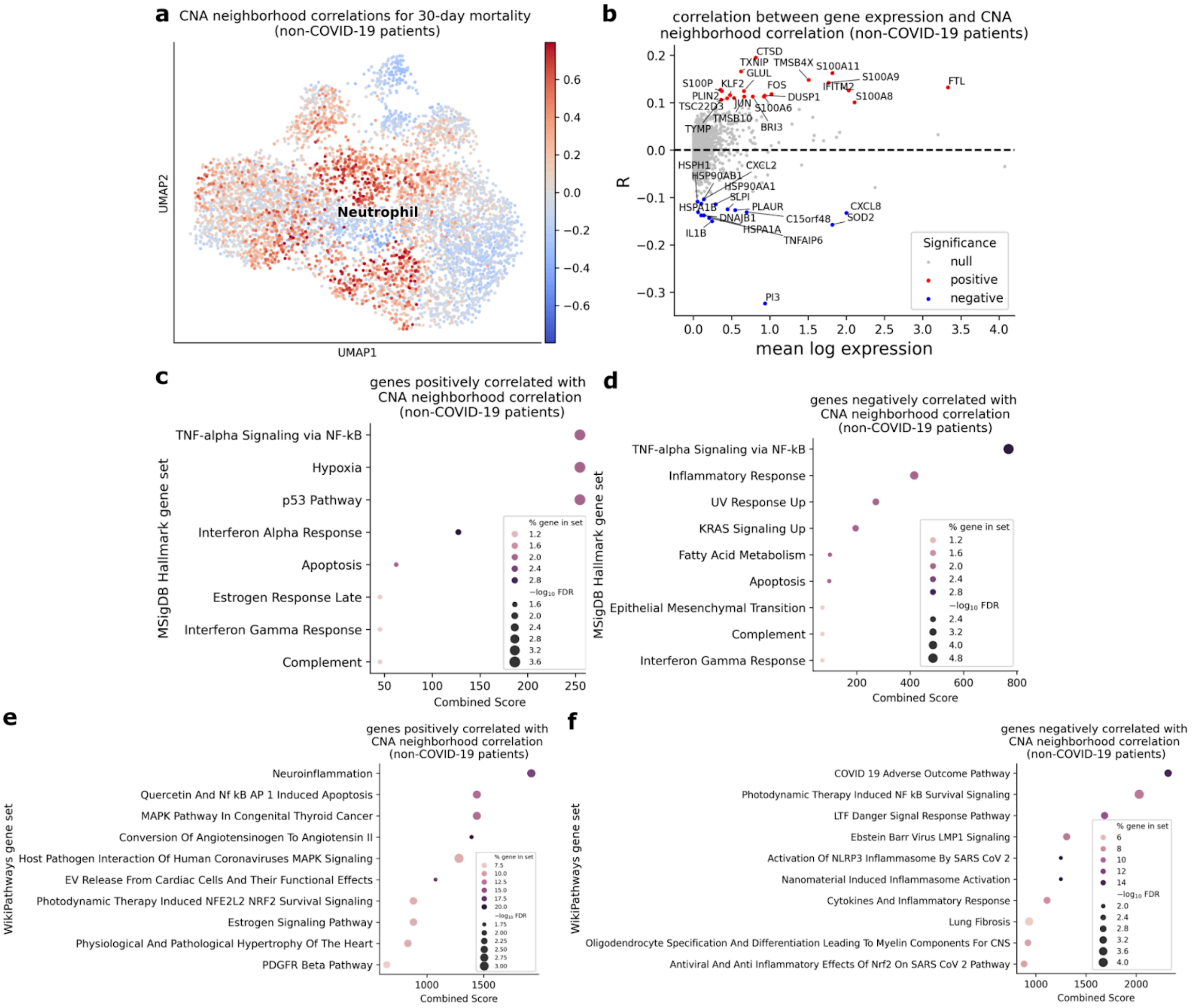


Supplementary Figure 5: **Results from the neutrophil-focused CNA analysis of the COMET ETA scRNA-seq data using non-COVID-19 patients.** (**a**) We report the CNA neighborhood correlations for 30-day mortality obtained using non-COVID-19 patients only. (**b**) We report the correlation between gene expression and CNA neighborhood correlation. Significant correlations (FDR < 0.05, |R| > 0.1) are highlighted in colors. (**c**, **d**) We report the MSigDB Hallmark gene sets significantly (FDR < 0.05) overlapping genes positively and negatively correlated with CNA neighborhood correlations, respectively.

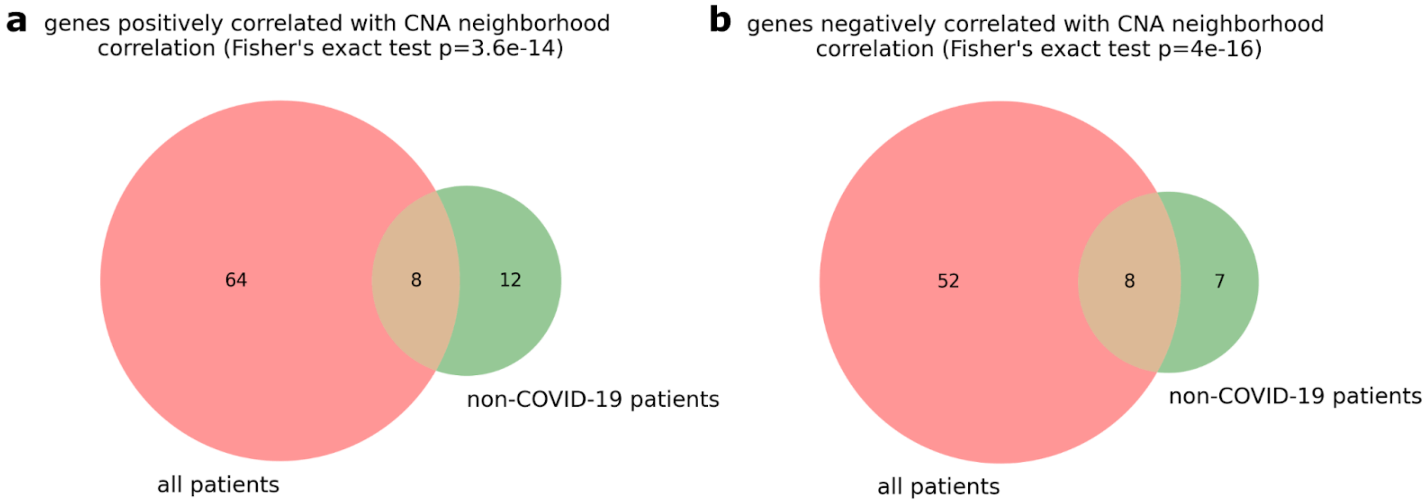


Supplementary Figure 6: **Overlap between genes significantly correlated with CNA neighborhood correlations in analysis of neutrophils of all and non-COVID-19 patients.** (**a**) We report the overlap of genes significantly positively (FDR < 0.05, R > 0.1) correlated with CNA neighborhood correlation between analyses of all vs. non-COVID-19 patients. (**b**) We report the overlap of genes significantly negatively (FDR < 0.05, R < -0.1) correlated with CNA neighborhood correlation between analyses of all vs. non-COVID-19 patients. P-values testing the significance of overlap were obtained using Fisher’s exact test.

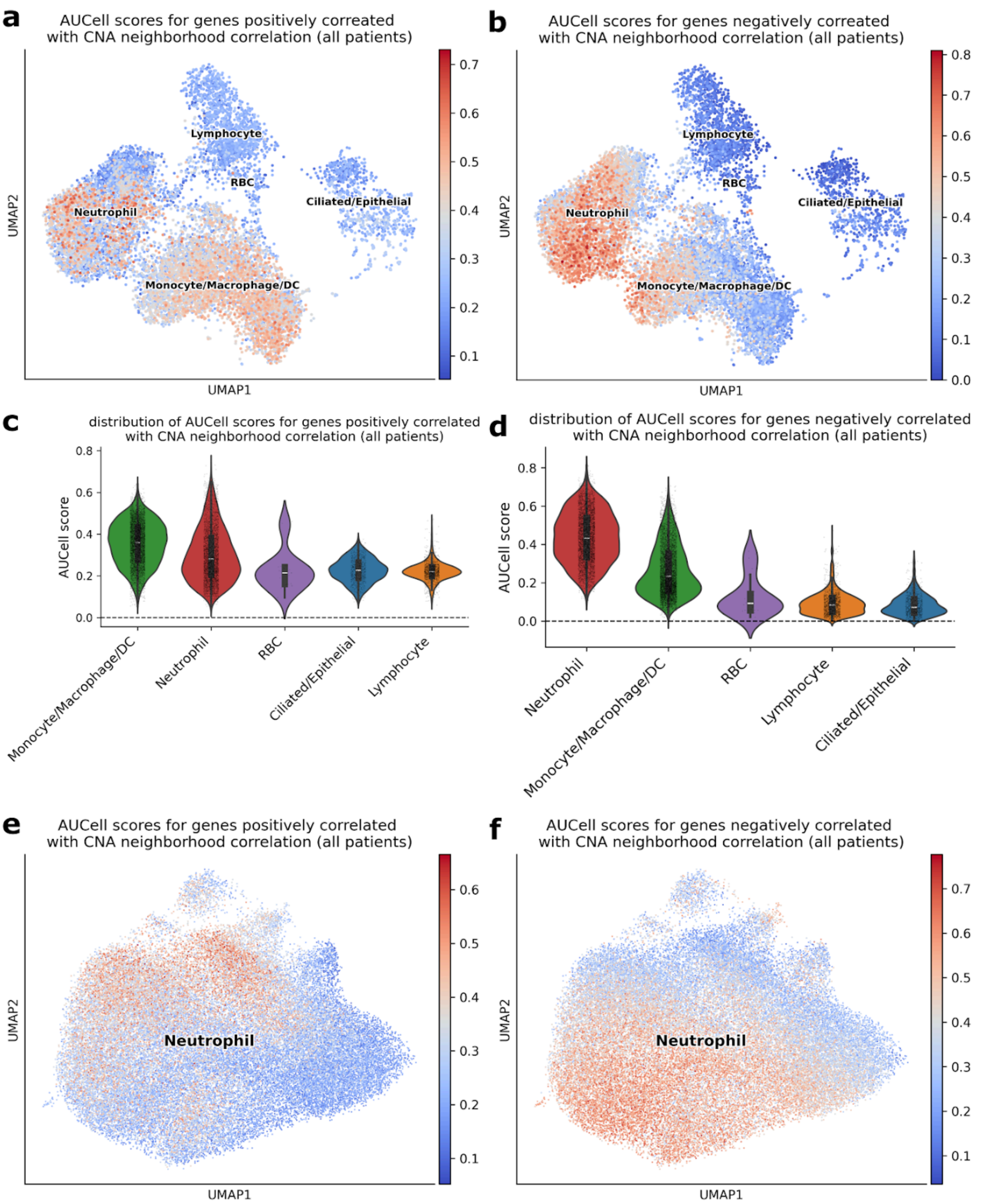


Supplementary Figure 7: **AUCell scores of the CNA mortality-associated genes in the COMET scRNA-seq data.** (**a**, **b**) UMAP plots showing the AUCell scores calculated using CNA mortality-increasing and mortality-decreasing genes, respectively, across all major cell types in the COMET ETA scRNA-seq data. (**c**, **d**) Violin plots showing the distribution of AUCell scores calculated using CNA mortality-increasing and mortality-decreasing genes, respectively, across the major cell types in the COMET ETA scRNA-seq data. The white lines in the violin plots represent the median. (**e**, **f**) UMAP plots showing the AUCell scores calculated using CNA mortality-increasing and mortality-decreasing genes, respectively, across the neutrophils in the COMET ETA scRNA-seq data.

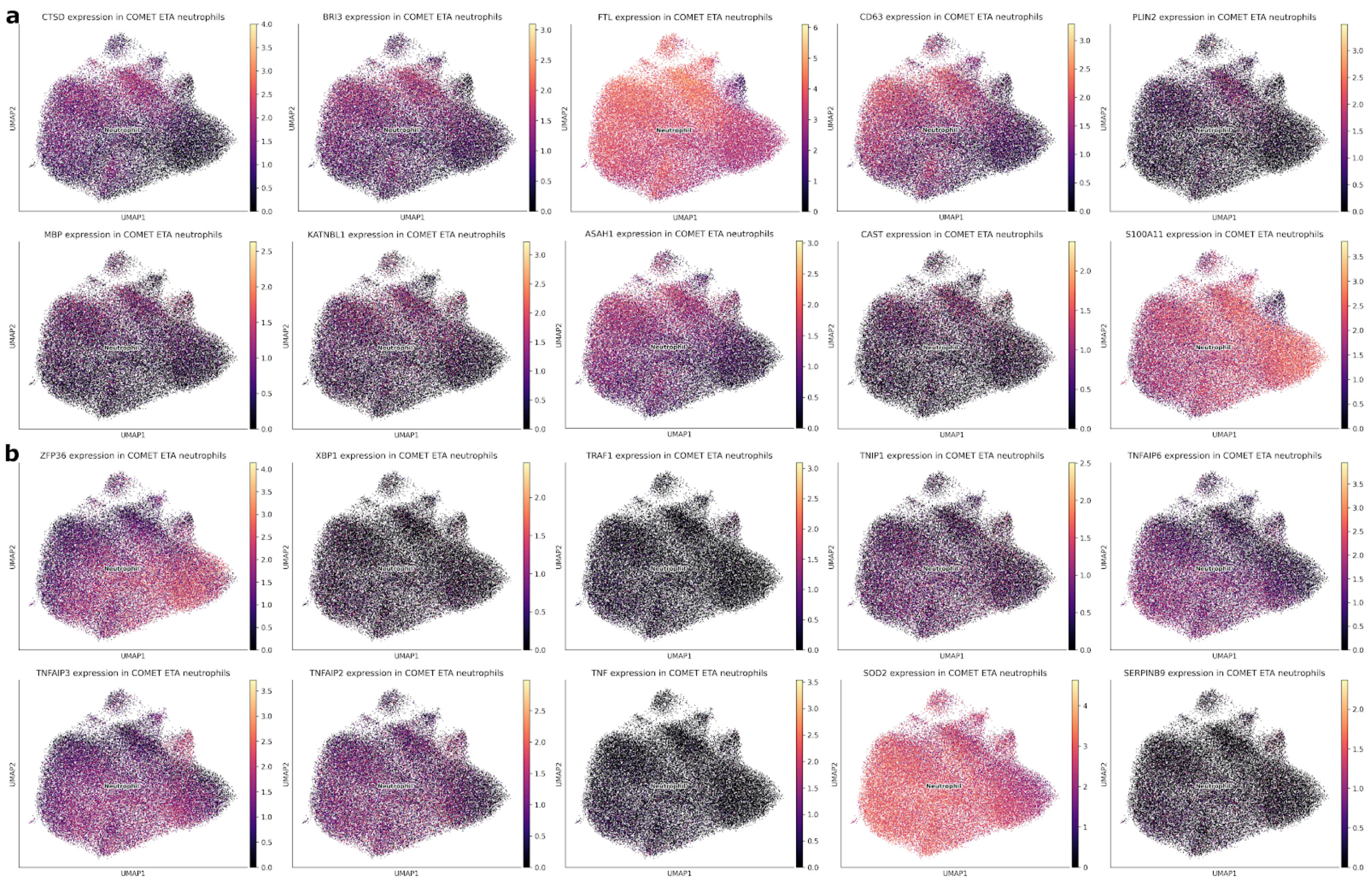


Supplementary Figure 8: **Expression of the CNA mortality-associated genes in COMET ETA neutrophils.** UMAP plots showing the expression of top 10 genes whose expression most positively (**a**) and negatively (**b**) correlated (|R| > 0.1, FDR < 0.05) with CNA neighborhood correlation for 30-day mortality in COMET ETA neutrophils.

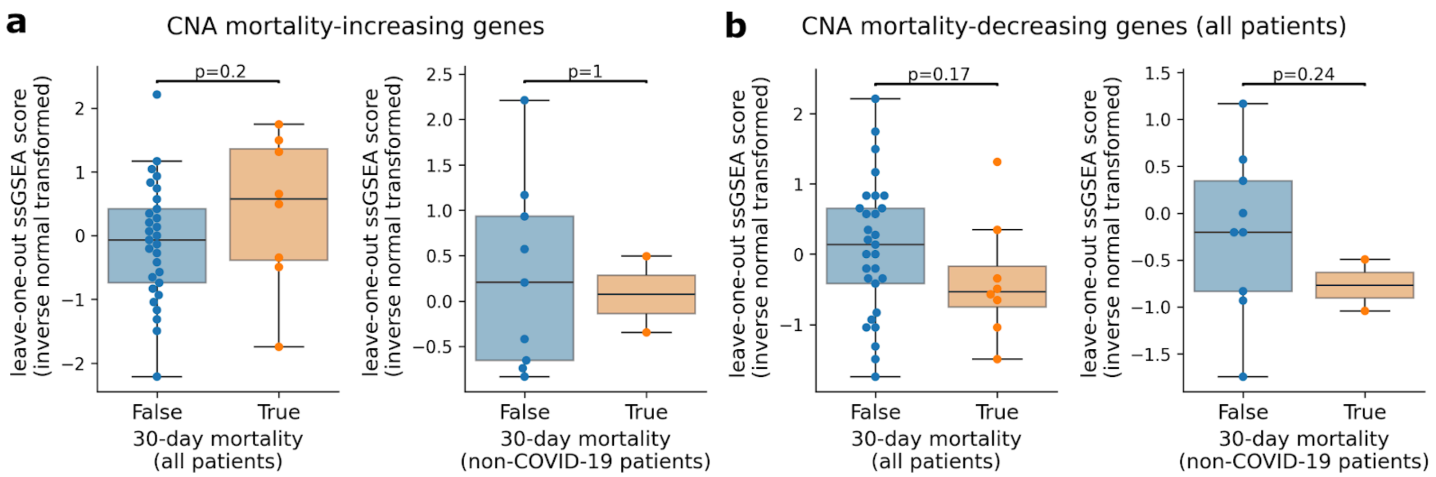


Supplementary Figure 9: **Median leave-one-out AUCell scores of the CNA mortality-associated genes across neutrophils of all patients.** (**a**) We report median leave-one-out AUCell scores, calculated using CNA mortality-increasing genes, across neutrophils of all patients. (**b**) We report median leave-one-out AUCell scores, calculated using CNA mortality-decreasing genes, across neutrophils of all patients. The dark lines in the box plots represent the median, with the boxes representing the interquartile range (IQR), and whiskers representing 1.5× IQR. P-values were obtained using Student's t-test with 1 degree of freedom.

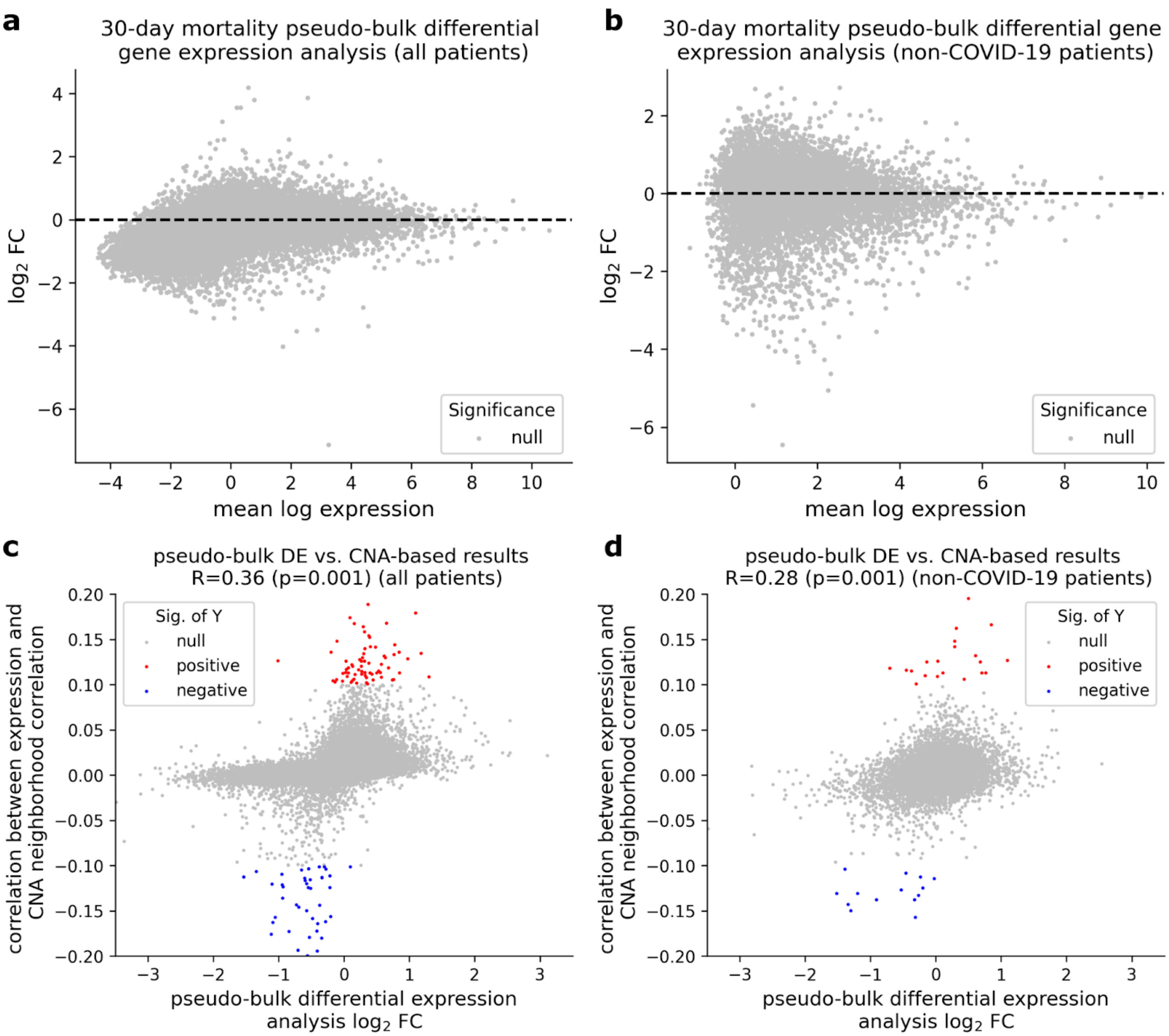


Supplementary Figure 10: **Comparison between pseudobulk differential expression and CNA analysis** (**a**, **b**) MA plots showing the results of pseudobulk gene expression using all patients and non-COVID-19 patients, respectively. (**c**, **d**) Comparison between $\log_2$ fold change from pseudobulk differential expression analysis (x-axis) and correlation between gene expression and CNA neighborhood correlation (y-axis) using all patients and non-COVID-19 patients, respectively. P-values were obtained based on 1,000 permutations.

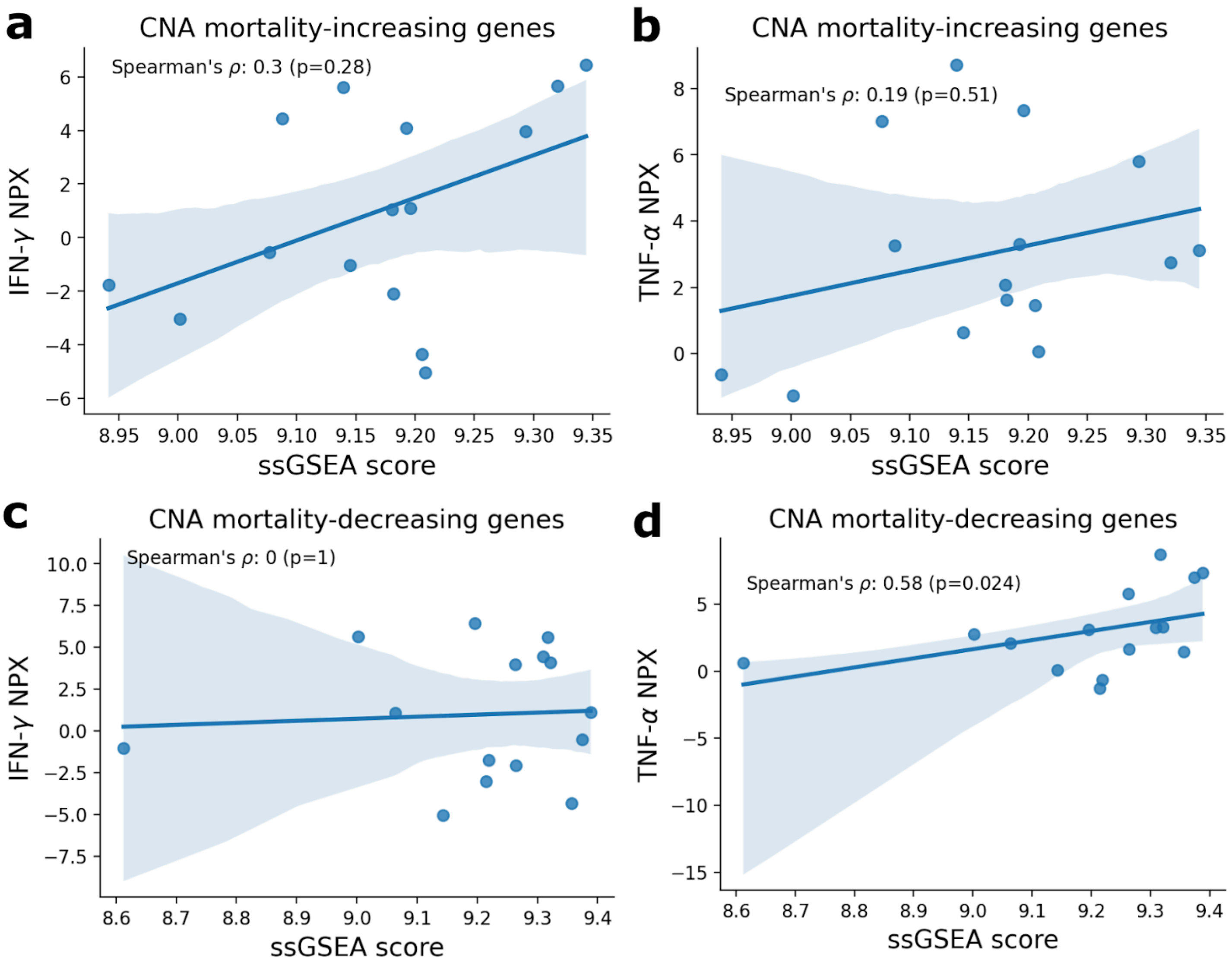


Supplementary Figure 11: **Association between the CNA mortality-associated gene signature scores and protein expression of IFN-ɣ and TNF-ɑ in ETA.** (**a**, **b**) We report the Spearman's correlation between ssGSEA scores calculated using the CNA mortality-increasing genes in airway neutrophils and the protein levels of IFN-ɣ and TNF-ɑ in ETA, respectively. (**c**, **d**) We report the Spearman's correlation between ssGSEA scores calculated using the CNA mortality-decreasing genes and the protein expression of IFN-γ and TNF-α, respectively. All ssGSEA scores were calculated based on the pseudobulked gene expression of neutrophils in the COMET ETA scRNA-seq data. Each dot in the scatter dot represents a patient, for whom both ETA scRNA-seq and O-link protein expression data were available; solid lines represent the regression line, with shaded regions representing the 95% confidence intervals. Two-tailed p-values testing the significance of the Spearman's correlation were based on 1,000 permutations.

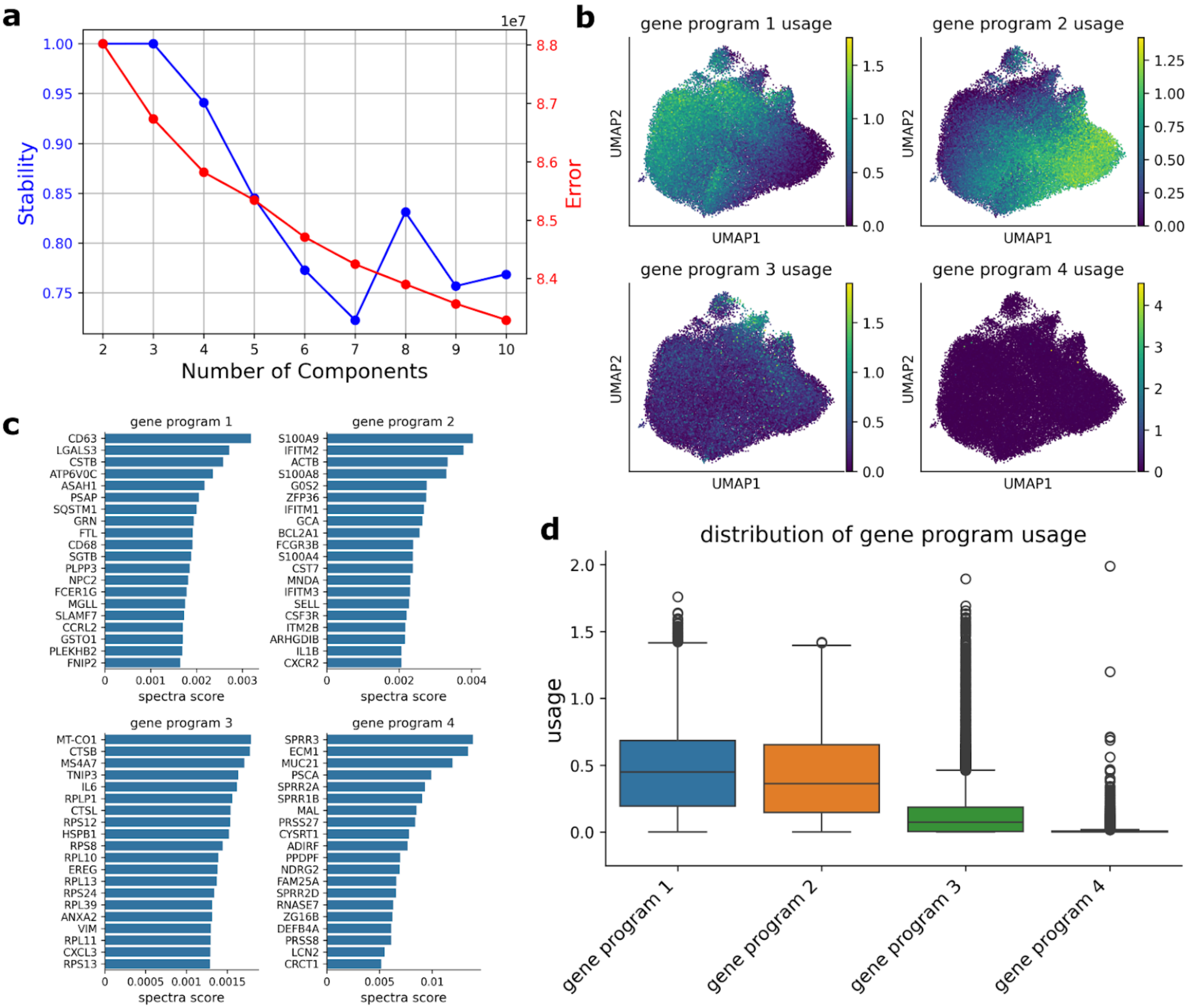


Supplementary Figure 12: **Results from the cNMF gene program analysis of neutrophils in the COMET ETA scRNA-seq data.** (**a**) k selection plot for the cNMF analysis. (**b**) UMAP plots showing the usage of each of the 4 inferred gene programs. (**c**) Top 20 genes with the highest spectra score for each of the 4 gene programs. (**d**) Distribution of usage for each of the 4 inferred gene programs. The dark lines in the box plots represent the median, with the boxes representing the interquartile range (IQR), and whiskers representing 1.5× IQR.

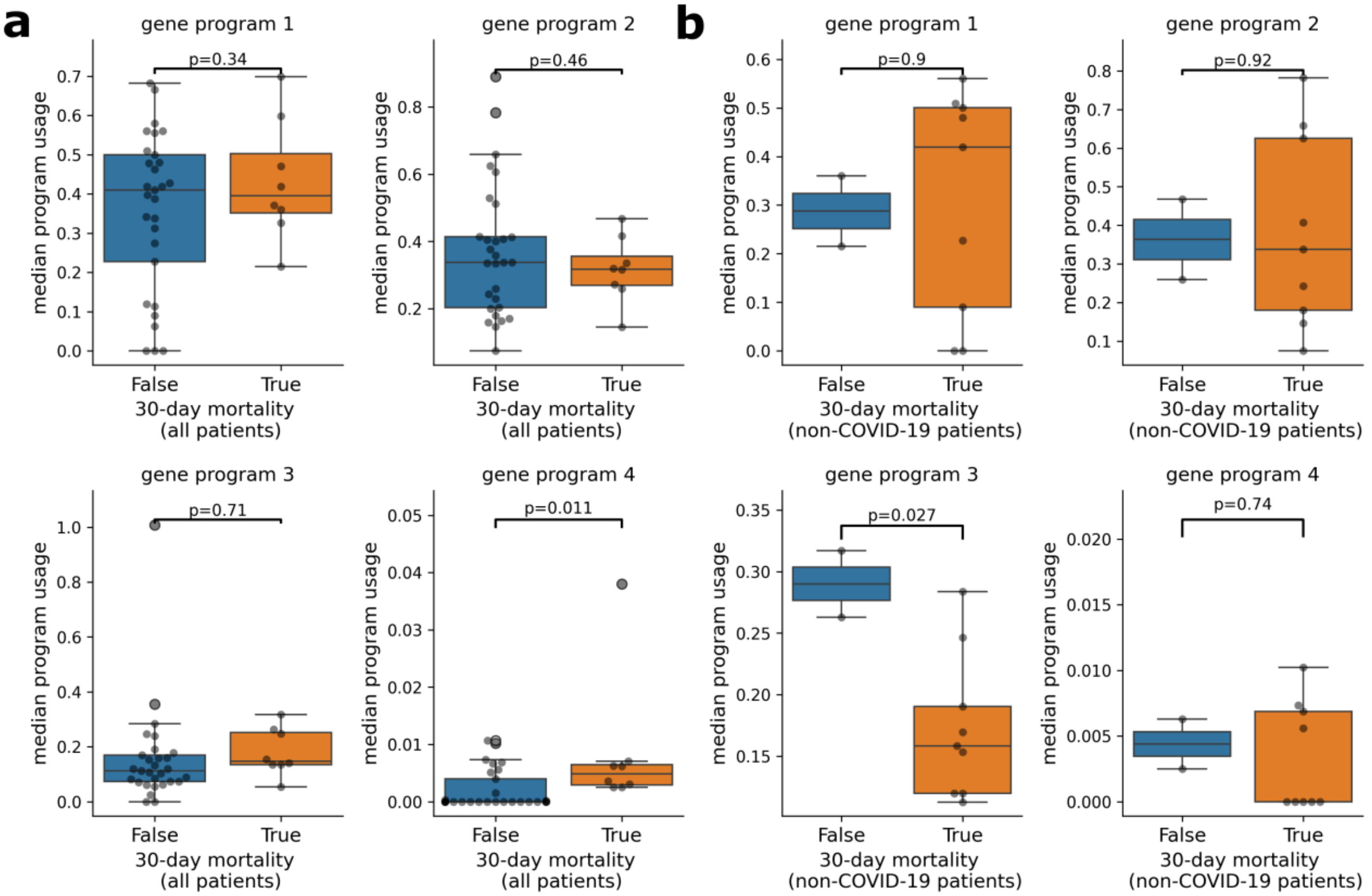


Supplementary Figure 13: **Association between 30-day mortality and cNMF neutrophil gene program.** (**a**) We report the association between 30-day mortality and median usage of each gene program across all patients. (**b**) We report the association between 30-day mortality and median usage of each gene program across non-COVID-19 patients. Each dot in the box plots represents a patient. The dark lines in the box plots represent the median, with the boxes representing the interquartile range (IQR), and whiskers representing 1.5× IQR. Two-tailed p-values testing the difference  were obtained using Student's t-test with 1 degree of freedom.

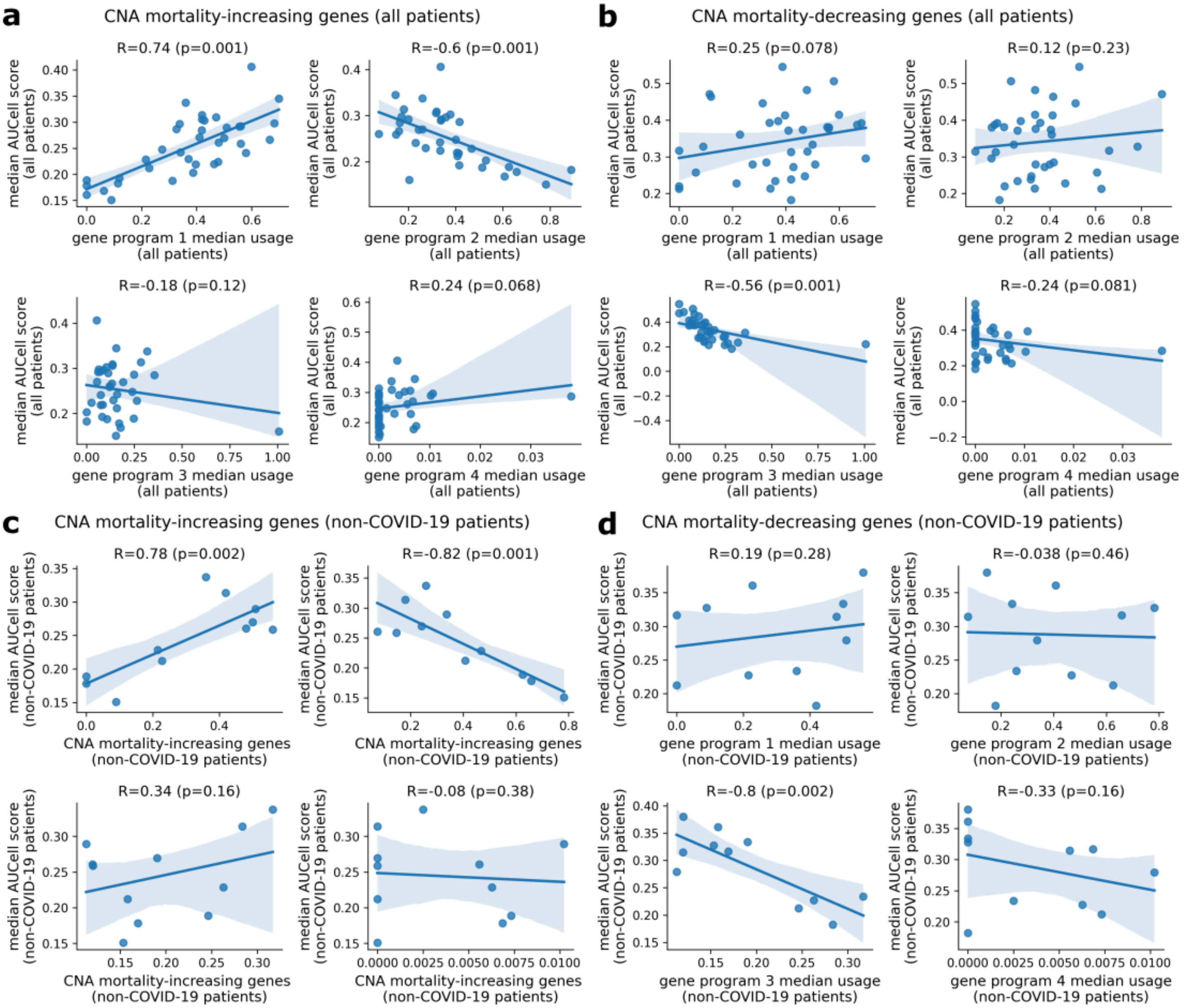


Supplementary Figure 14: **Association between cNMF gene program usage and AUCell scores calculated using CNA mortality-associated genes.** (**a**, **b**) We report the correlation between the median gene program usage and AUCell scores calculated using mortality-increasing and decreasing genes, respectively, across all patients. (**c**, **d**) We report the correlation between the median gene program usage and AUCell scores calculated using mortality-increasing and decreasing genes, respectively, across non-COVID-19 patients. Each dot in the scatter plot represents a patient in the COMET ETA scRNA-seq data; solid line represents the regression slope; error bars represent 95% confidence intervals.

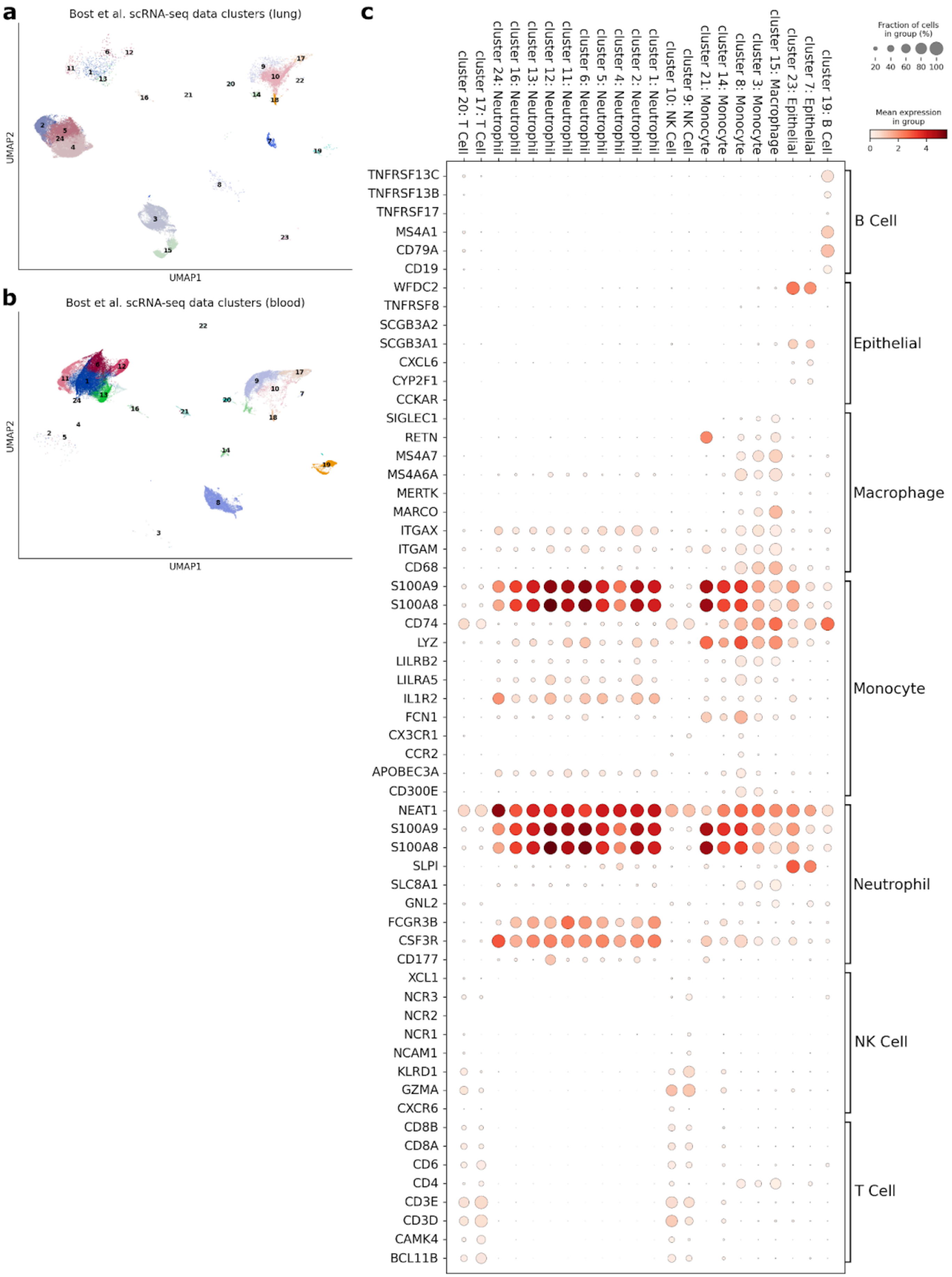
a
Bost et al. scRNA-seq data clusters (lung)
UMAP2
UMAP1
b
Bost et al. scRNA-seq data clusters (blood)
UMAP2
UMAP1
c
cluster 20: T Cell
cluster 17: T Cell
cluster 24: Neutrophil
cluster 16: Neutrophil
cluster 13: Neutrophil
cluster 12: Neutrophil
cluster 11: Neutrophil
cluster 6: Neutrophil
cluster 5: Neutrophil
cluster 4: Neutrophil
cluster 2: Neutrophil
cluster 1: Neutrophil
cluster 10: NK Cell
cluster 9: NK Cell
cluster 21: Monocyte
cluster 14: Monocyte
cluster 8: Monocyte
cluster 3: Monocyte
cluster 15: Macrophage
cluster 23: Epithelial
cluster 7: Epithelial
cluster 19: B Cell
Fraction of cells in group (%)
20 40 60 80 100
Mean expression in group
0 2 4
TNFRSF13C
TNFRSF13B
TNFRSF17
MS4A1
CD79A
CD19
WFDC2
TNFRSF8
SCGB3A2
SCGB3A1
CXCL6
CYP2F1
CCKAR
SIGLEC1
RETN
MS4A7
MS4A6A
MERTK
MARCO
ITGAX
ITGAM
CD68
S100A9
S100A8
CD74
LYZ
LILRB2
LILRA5
IL1R2
FCN1
CX3CR1
CCR2
APOBEC3A
CD300E
NEAT1
S100A9
S100A8
SLPI
SLC8A1
GNL2
FCGR3B
CSF3R
CD177
XCL1
NCR3
NCR2
NCR1
NCAM1
KLRD1
GZMA
CXCR6
CD8B
CD8A
CD6
CD4
CD3E
CD3D
CAMK4
BCL11B
B Cell
Epithelial
Macrophage
Monocyte
Neutrophil
NK Cell
T Cell

Supplementary Figure 15: **Annotating the Bost et al. COVID-19 scRNA-seq data**. (**a**, **b**) UMAP plot showing clusters obtained for the Bost et al. lung and blood scRNA-seq data, respectively (see Methods for the details of the clustering procedure). (**c**) Dot plot of mean expression and fraction of cells expressing for each (gene, cluster) pair, with clusters (and annotated cell types) indicated in columns, and genes indicated by rows. Groups of genes used as markers for particular cell types are indicated by brackets on the right-hand side.

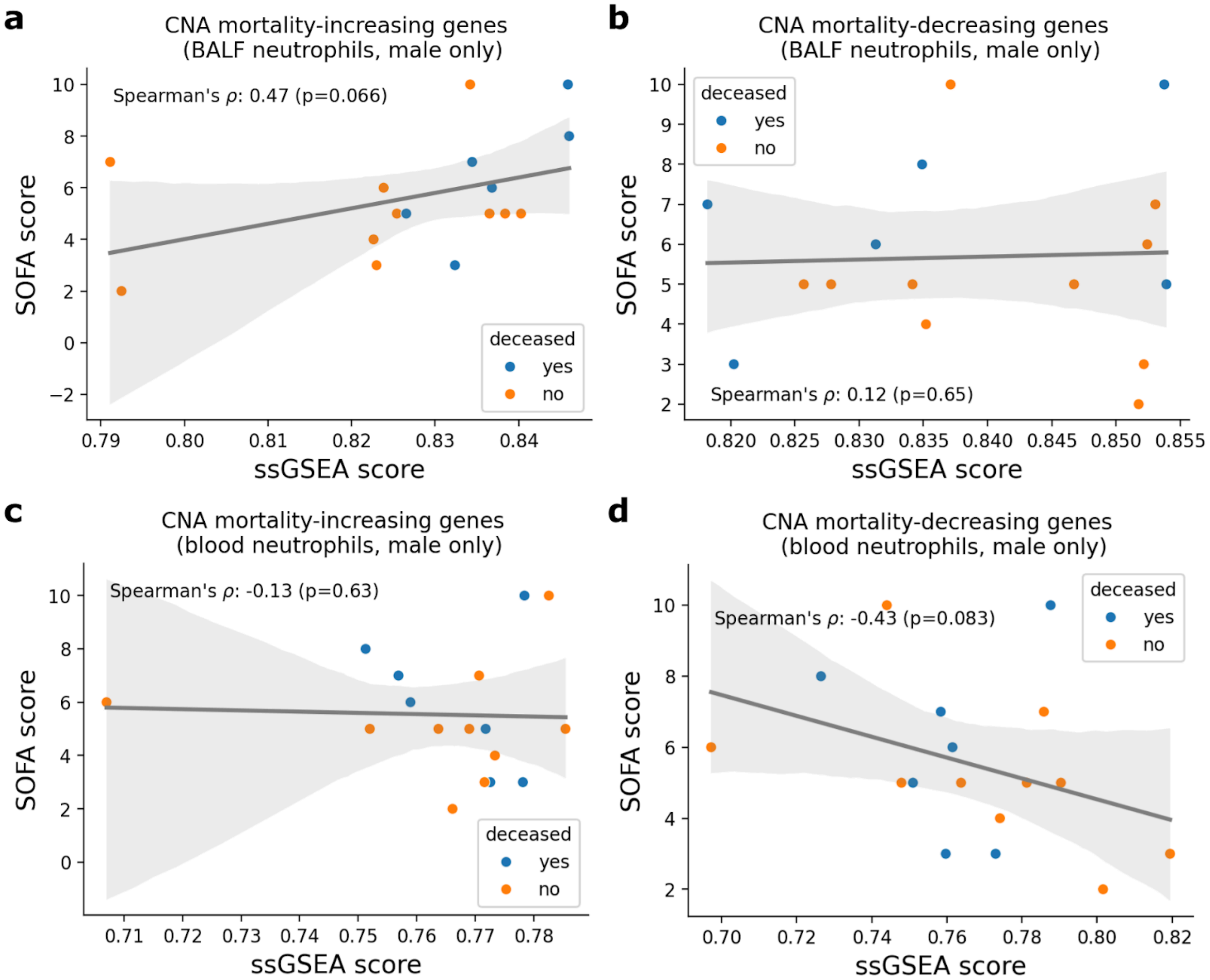


Supplementary Figure 16: **Results from the analysis of CNA mortality-associated genes in the neutrophils from the Bost et al. male COVID-19 patient scRNA-seq data.** (**a**, **b**) We report the Spearman's correlation between patient SOFA scores and ssGSEA scores for CNA mortality-increasing and decreasing genes, respectively, for BALF; we report analogous results for blood neutrophils in (**c**, **d**). ssGSEA scores were calculated based on log-normalized pseudobulk gene expression. Solid lines in the scatter plots are regression lines fitting the SOFA scores against the ssGSEA scores; shaded regions represent 95% confidence intervals.

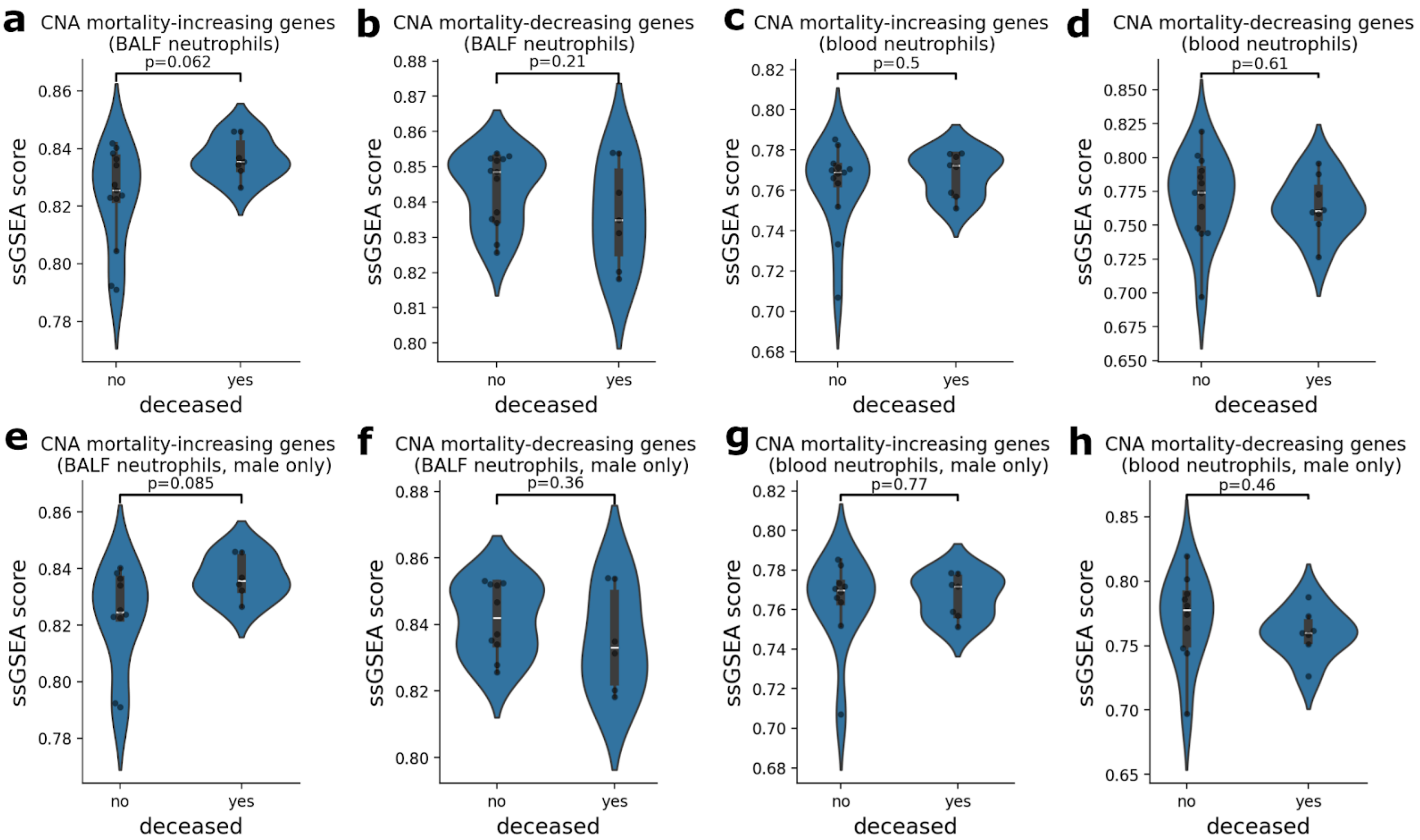


Supplementary Figure 17: **Associations between mortality and the neutrophil-specific ssGSEA scores calculated using the CNA mortality-associated genes in the Bost et al. COVID-19 scRNA-seq data.** (**a**, **b**) We report the association between mortality status and neutrophil-specific ssGSEA scores calculated using the CNA mortality-increasing and decreasing genes, respectively, using the BALF scRNA-seq data; analogous results for CNA mortality-increasing and decreasing genes obtained using the blood neutrophils are reported in (**c**, **d**), respectively. (**e**, **f**) We report the association between mortality status and neutrophil-specific ssGSEA scores calculated using the CNA mortality-increasing and decreasing genes, respectively, using the BALF scRNA-seq data from male donors; analogous results for CNA mortality-increasing and decreasing genes obtained using the blood neutrophils are reported in (**g**, **h**). The white lines in the violin plots represent the median. Each dot in the violin plots represents a donor; two-tailed p-values were obtained based on 1,000 permutations.

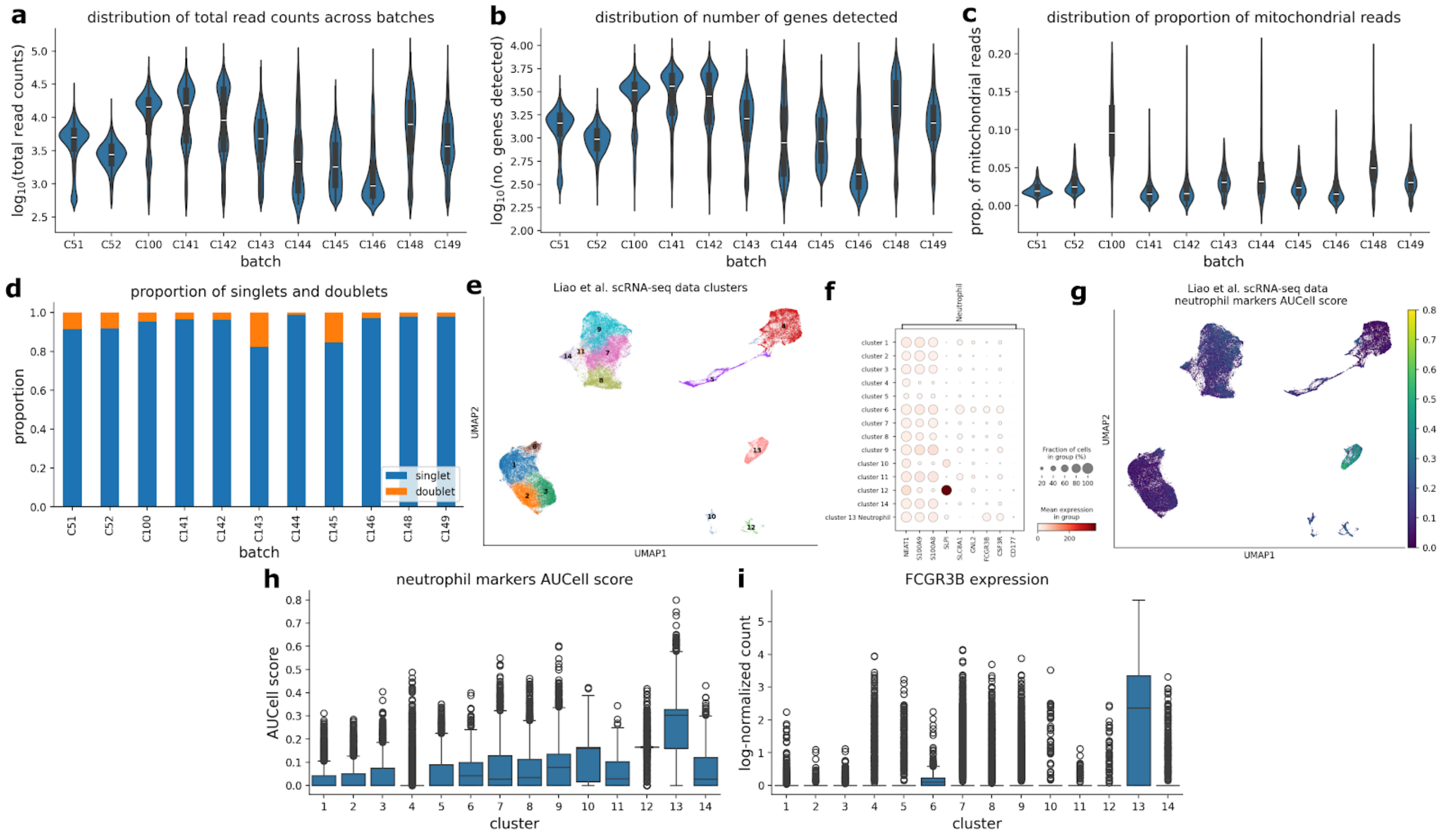


Supplementary Figure 18: **Preprocessing and identification of neutrophils for the Liao et al. COVID-19 BALF scRNA-seq data.** (**a**, **b**, **c**) We report the distribution of log10 total read counts, log10 number of genes detected, and proportion of mitochondrial reads, respectively, for cells in each batch. (**d**) We report the proportions of singlets and doublets detected in each batch. (**e**) UMAP plots showing the clusters detected in the scRNA-seq data (see Methods for details of the clustering). (**f**) Dot plot of mean expression and fraction of cells expressing for each (gene, cluster) pair, with clusters (and annotated cell types) indicated in columns, and genes indicated by rows. Groups of genes used as markers for neutrophils are indicated by brackets on the right-hand side. (**g**) UMAP plot showing the AUCell scores calculated using the neutrophil marker genes. (**h**) We report the distribution of AUCell scores calculated using the neutrophil maker genes for cells in each cluster. (**i**) We report the distribution of the expression of *FCGR3B* in each cluster. The white lines in the violin plots represent the median. The dark lines in the box plots represent the median, with the boxes representing the interquartile range (IQR), and whiskers representing 1.5× IQR.

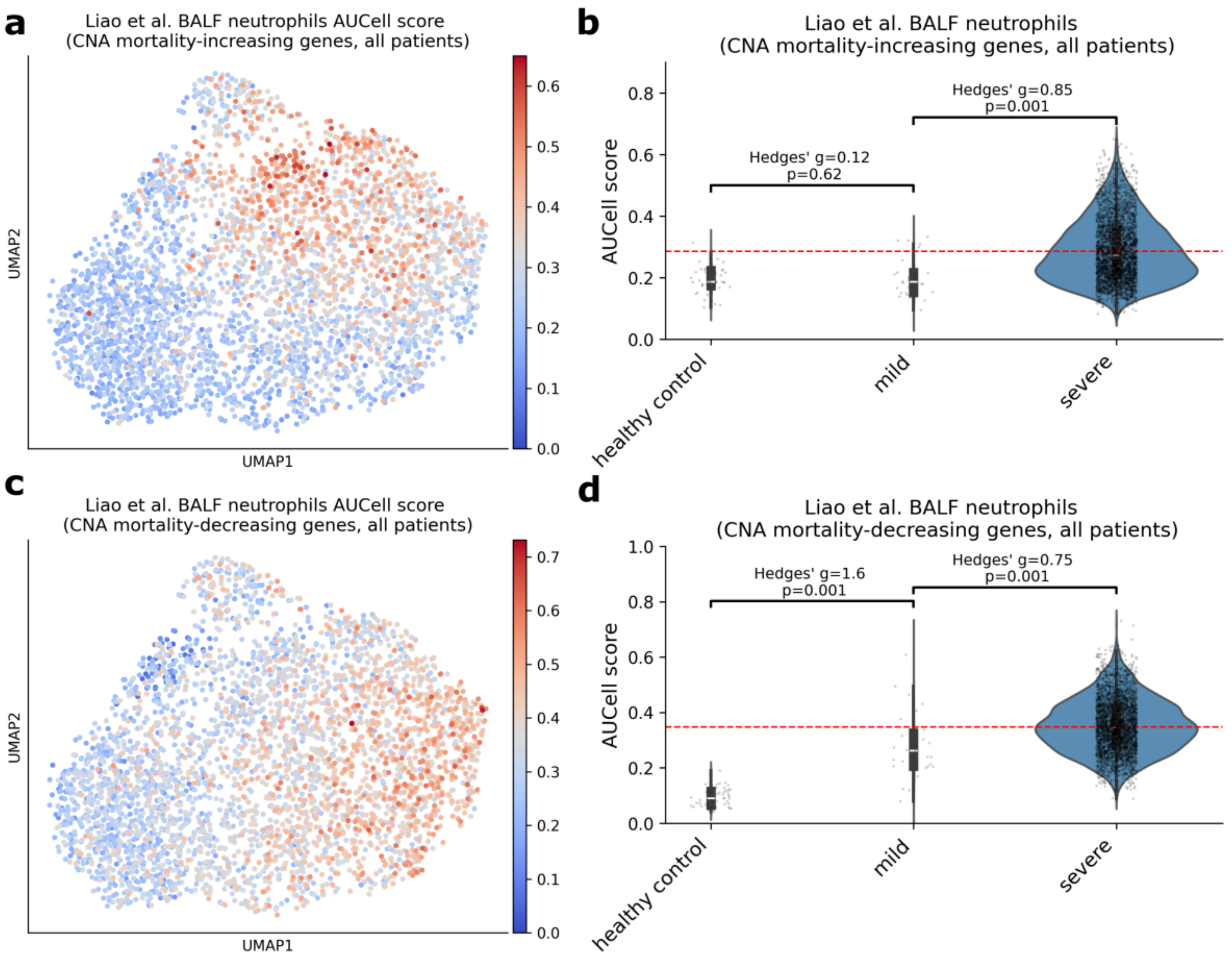


Supplementary Figure 19: **Results from the analysis of CNA mortality-associated genes using the neutrophils from the Liao et al. COVID-19 BALF scRNA-seq data.** (**a**, **c**) UMAP plot showing the AUCell scores calculated using the CNA mortality-increasing and decreasing genes, respectively, for the BALF neutrophils. (**b**, **d**) We report the distributions of AUCell scores calculated using the CNA mortality-increasing and decreasing genes, respectively, across the BALF neutrophils from donors of varying degrees of disease severity. The white lines in the violin plots represent the median. Red dashed lines represent the mean AUCell scores across all neutrophils. Two-tailed p-values testing the statistical significance of the difference in mean AUCell scores across different severity were obtained based on 1,000 permutations.

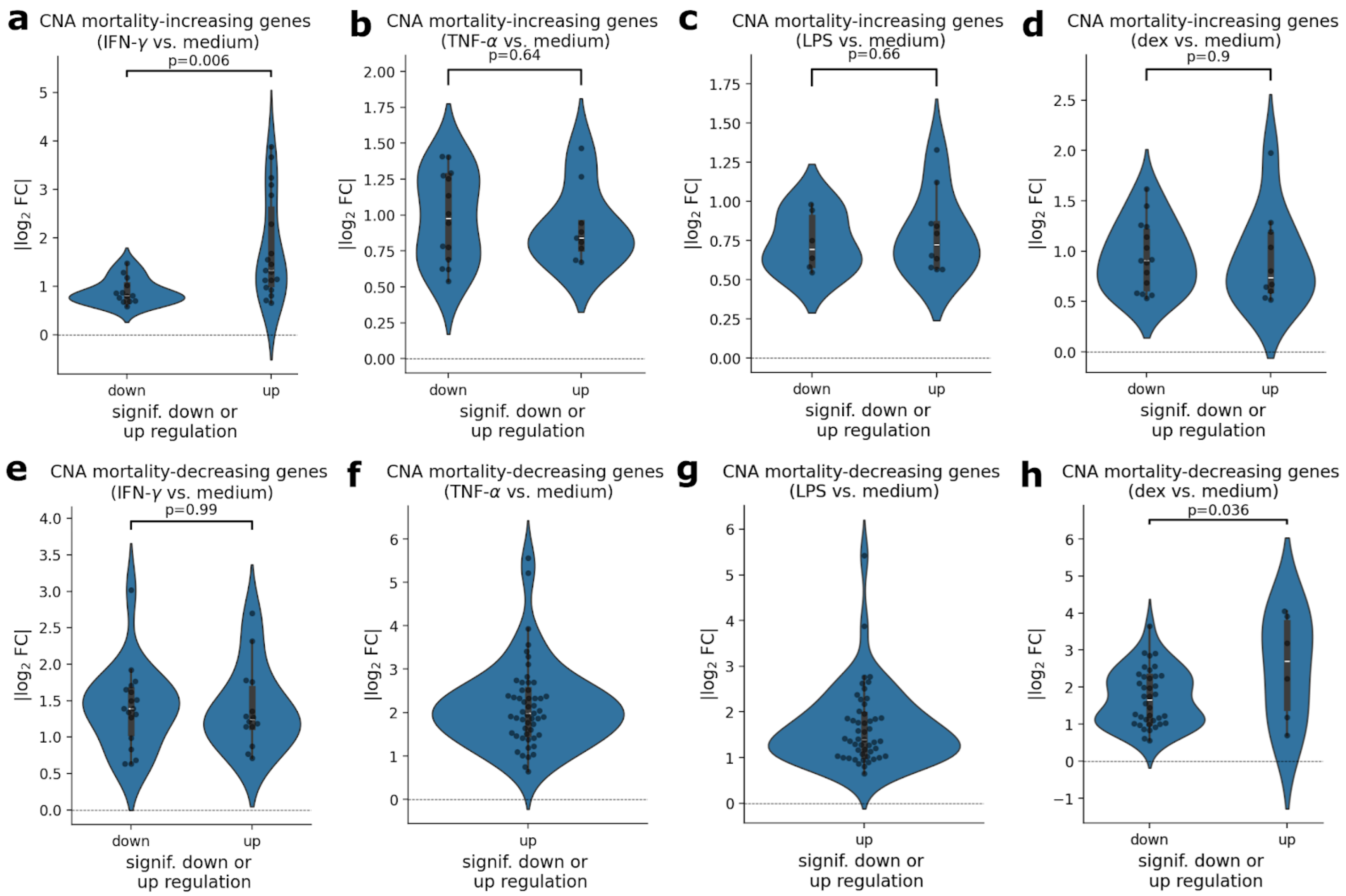


Supplementary Figure 20: **Distributions of |log$_2$ FC| of CNA mortality-associated genes in human blood neutrophils *in vitro* experiments.** (**a, b, c, d**) We report the distributions of |log$_2$ FC| of CNA mortality-increasing genes in DE analyses (relative to medium) of neutrophils stimulated with IFN-γ, TNF-α, lipopolysaccharide (LPS), and dexamethasone (dex), respectively. We report analogous results for CNA mortality-decreasing genes in (**e**, **f**, **g**, **h**). The white lines in the violin plots represent the median.

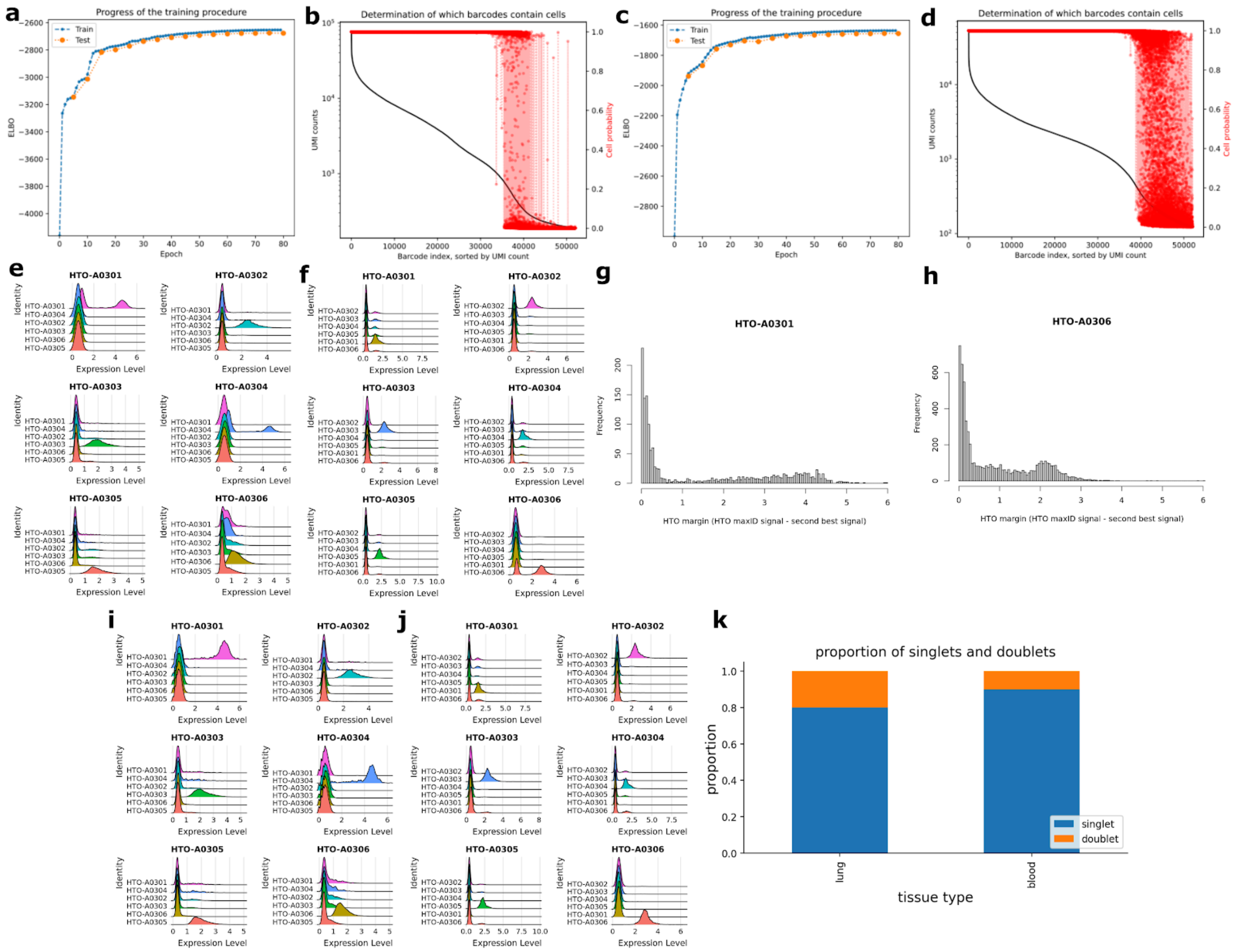


Supplementary Figure 21: **Preprocessing of the mouse flu model scRNA-seq data**. (**a**, **c**) Training of CellBender models of ambient + “true” RNA expression for lung (**a**) and blood (**c**) sequencing pools. (**b**, **d**) Droplet barcode indices sorted by UMI count for lung and blood, respectively, with UMI count delineated by black curve, and cell probability by red dot. All droplets with probability > 0.5 are accepted as probable cells for subsequent processing. (**e**, **f**): Assignment of droplets to sample pools based on HTO barcode counts for lung (**e**) and blood (**f**). (**g**, **h**): Distribution of difference between max HTO signal and next best signal (HTO margin) for problematic samples in lung (**g**) and blood (**h**). (**i**, **j**) Sample pool assignment after removal of low-quality droplets with mixed HTO identity (i.e. HTO margin < 0.5), for lung (**i**), and blood (**j**). (**k**) Proportions of singlets and doublets detected in the scRNA-seq data in lung and blood.

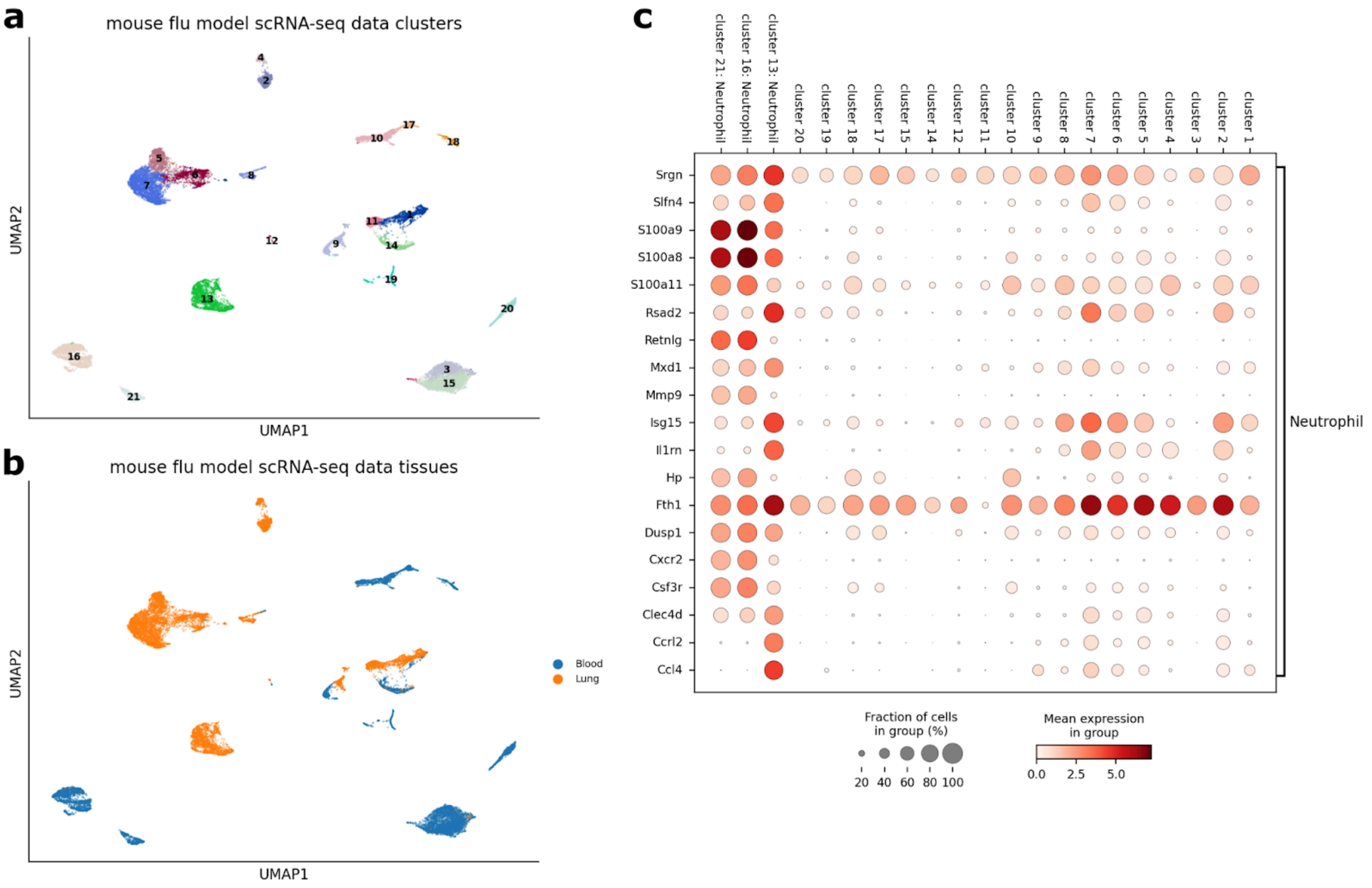


Supplementary Figure 22: **identification of neutrophils in the mouse flu model scRNA-seq data.** (**a**) UMAP plot showing the clusters defined for the mouse flu model scRNA-seq data. (**b**) UMAP plot showing cells from blood and lung. (**c**) Dot plot of mean expression and fraction of cells expressing for each (gene, cluster) pair, with clusters (and annotated cell types) indicated in columns, and genes indicated by rows. Groups of genes used as markers for neutrophils are indicated by brackets on the right-hand side.

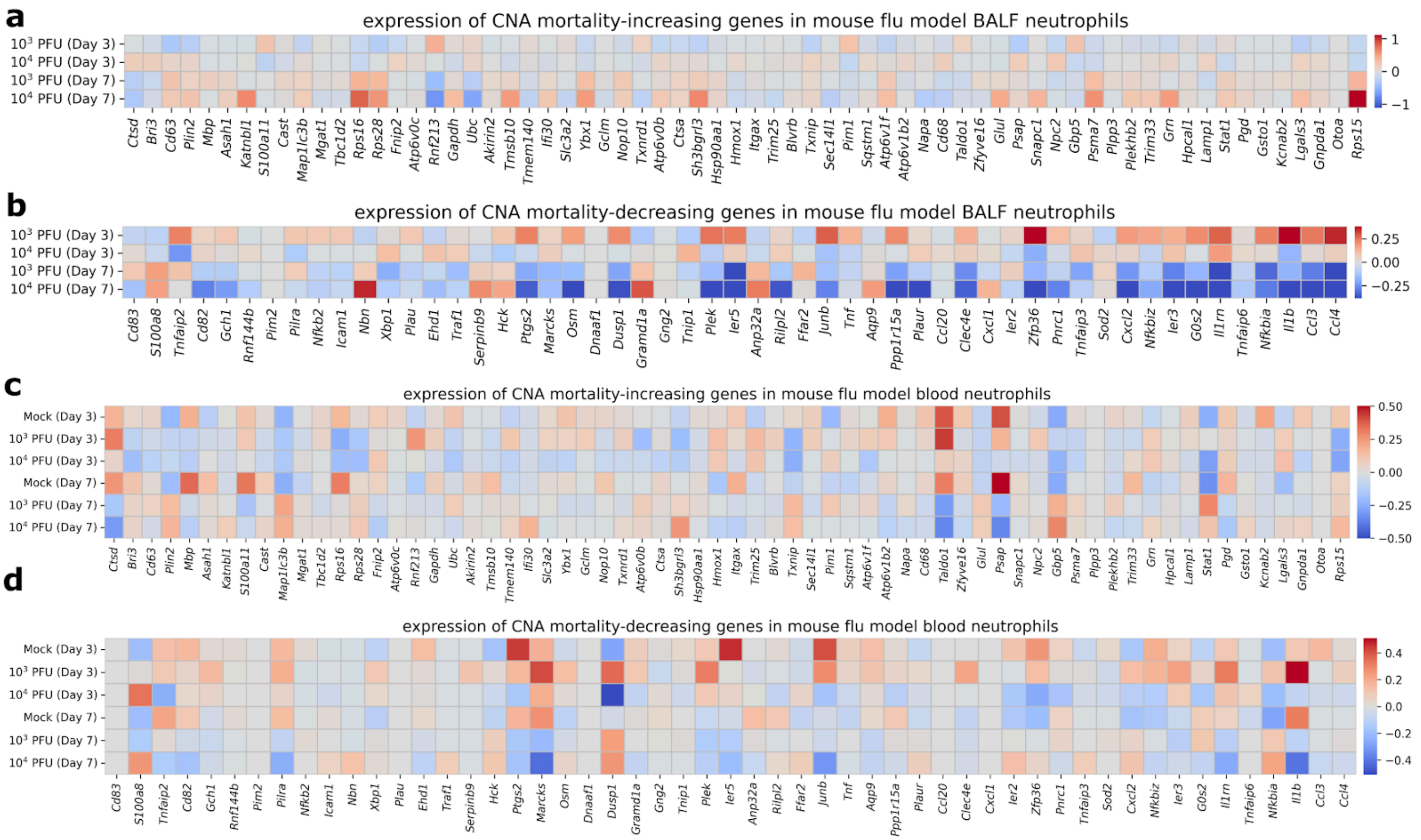


Supplementary Figure 23: **Expression of CNA mortality-associated genes in the mouse flu model scRNA-seq data.** (**a**, **b**) We report the mean expression of the CNA mortality-increasing and decreasing genes, respectively, for the BALF neutrophils from the mouse flu model scRNA-seq data. Analogous results for the blood neutrophils from the mouse flu model scRNA-seq data (**c**, **d**).

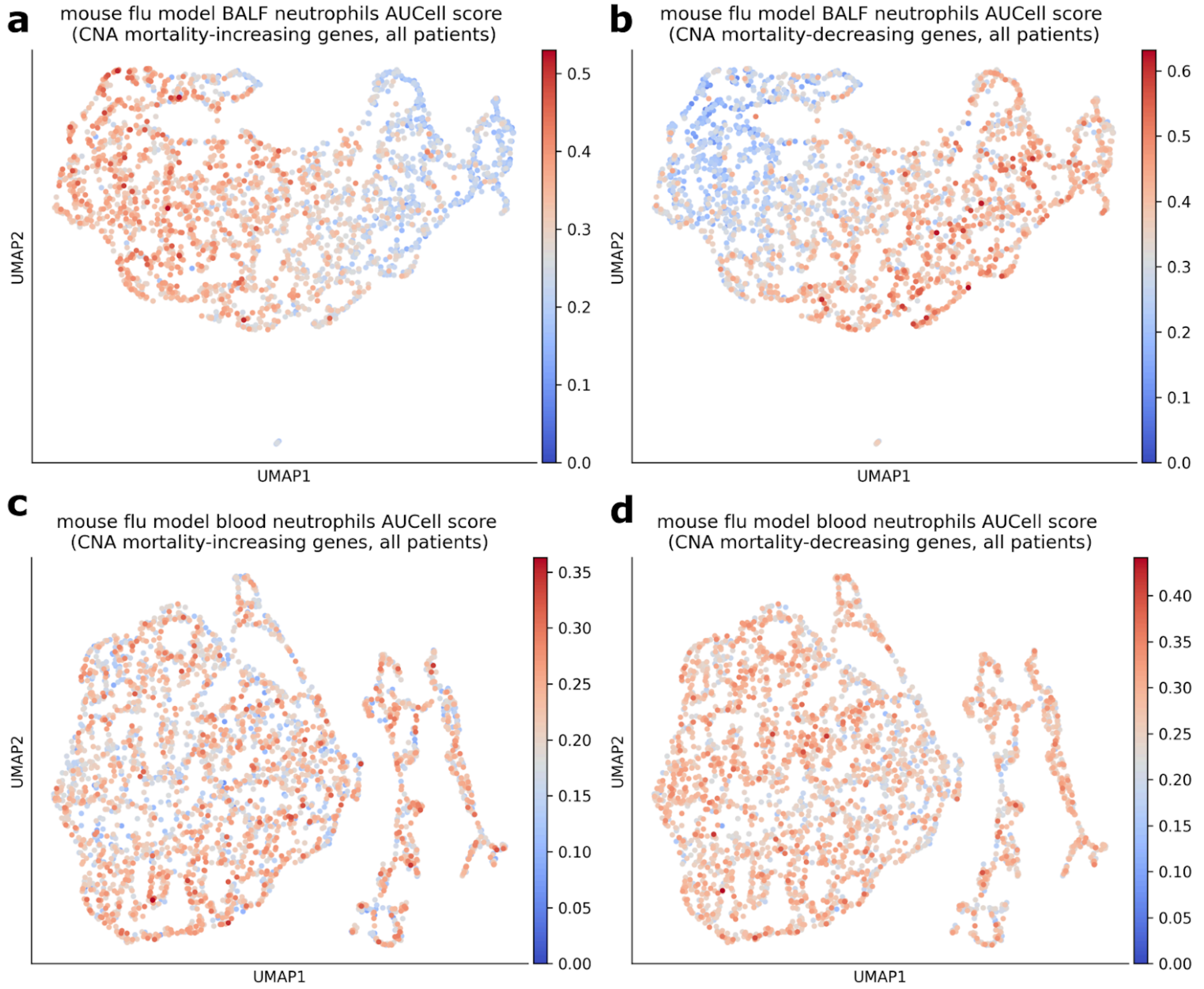


Supplementary Figure 24: **Association between flu virus dosage and neutrophil-specific AUCell scores for CNA mortality-associated genes in the mouse flu model scRNA-seq data.** (**a, b**) UMAP plot showing the AUCell scores calculated using the CNA mortality-increasing and decreasing genes, respectively, for the BALF neutrophils; analogous results for blood neutrophils are reported in (**c**, **d**).
